# Dual Targeting of DNA and EGFR by ZYH005 Induces DNA Damage and Mitotic Catastrophe in Glioblastoma

**DOI:** 10.1101/2025.04.14.25325787

**Authors:** Jianzheng Huang, Zijun Zhang, Yang Xiao, Ziming Zhao, Zengwei Luo, Junjun Liu, Suitian Lai, Chao Song, Shouchang Feng, Suojun Zhang, Xingjiang Yu, Qingyi Tong, Yonghui Zhang

## Abstract

**Background:** Glioblastoma (GBM) is an aggressive, therapy-resistant brain tumor with limited treatment options. Epidermal growth factor receptor (EGFR) is frequently amplified and activated in GBM, driving tumorigenesis through pro-oncogenic signaling and coordination of nuclear DNA repair. This study examines the anti-GBM efficacy and mechanism of ZYH005 (Z5), a brain-penetrant DNA intercalator exhibiting low systemic toxicity.

**Methods:** Antitumor efficacy of Z5 was determined with GBM cell lines and patient-derived glioblastoma stem cells (GSCs) in vitro and in vivo. Target identification and mechanistic validation were performed using DNA microarray, surface plasmon resonance, immunoblot and siRNA silencing. The EGFR-WEE1 correlation was analyzed via public databases and confirmed by coimmunoprecipitation.

**Results:** Z5 significantly inhibits the proliferation of GBM cell lines and patient-derived GSCs, effectively suppresses tumor growth in orthotopic GSC-induced mouse models, prolongs survival, and shows no obvious toxicity. Mechanistically, Z5 exerts potent anti-GBM activity through a dual mechanism: DNA intercalation-induced damage and targeted inhibition of EGFR. By specifically inhibiting EGFR at E762, Z5 not only enhances DNA damage by suppressing the DNA damage response in the nucleus but also disrupts the interaction between nuclear EGFR and WEE1, leading to impaired WEE1/CDC2 signaling and G2/M checkpoint failure. Extranuclearly, Z5 further enhances its anti-GBM efficacy by inhibiting the canonical EGFR downstream pathways, mTOR and ERK. Together, these molecular events promote cell cycle arrest and mitotic catastrophe in GBM cells.

**Conclusion:** Z5 emerges as a promising brain-penetrant clinical candidate for treating GBM. It acts through a dual mechanism that synergistically targets both DNA and EGFR, inducing mitotic catastrophe while demonstrating a favorable safety profile. These compelling findings provide a strong rationale for advancing Z5 toward clinical translation, offering a novel therapeutic strategy for GBM patients.

## 1. Background

Glioblastoma (GBM) stands as the most invasive and deadly primary brain tumor among adults[1]. It constitutes around 49% of all malignant central nervous system tumors. In high-income countries, its annual incidence is 3-7 cases per 100,000 individuals[2, 3]. The treatment of GBM adopts a multimodal strategy, integrating surgery, radiotherapy, and temozolomide (TMZ)-based chemotherapy. However, the median survival time for GBM patients is a mere 12-16 months, and the 5-year survival rate remains below 5%[2, 3]. This persistent therapeutic impasse underscores the critical need for paradigm-shifting treatment strategies in neuro-oncology.

Epidermal growth factor receptor (EGFR) and its constitutively active mutant EGFRvIII are well-established oncogenic drivers in GBM, with gene amplification occurring in over 50% of cases, particularly in the classical subtype[4]. Beyond activating canonical signaling pathways such as PI3K/AKT/mTOR, RAS/MAPK, and JAK/STAT—which collectively promote tumor proliferation, survival, and invasion—EGFR also translocates to the nucleus, where it directly facilitates DNA repair. There, it interacts with DNA-PKcs to enhance its phosphorylation, bolstering non-homologous end joining (NHEJ)[5, 6]. Furthermore, through the RAS/RAF/MEK/ERK and PI3K/AKT cascades, EGFR modulates the expression of DNA repair factors such as XRCC1 and RAD51, thereby enhancing DNA damage response (DDR) homeostasis and aiding tumor cells in evading genotoxic stress[7, 8]. This central role of EGFR in coordinating DDR underlies why its pharmacological inhibition not only disrupts DNA repair mechanisms but also sensitizes GBM cells to radiotherapy and DNA-damaging agents[9, 10]. Together, these multifaceted functions solidify EGFR’s position as a cornerstone therapeutic target in GBM.

Tyrosine kinase inhibitors (TKIs) show inhibitory activity against EGFR, their monotherapy efficacy in GBM is limited due to tumor heterogeneity, compensatory signaling pathways, and the blood–brain barrier (BBB)[11]. Accordingly, combination strategies—particularly pairing EGFR-TKIs with MEK/PI3K inhibitors or DNA-damaging agents (e.g., temozolomide, radiotherapy)—are required to achieve superior therapeutic outcomes. Preclinical studies further demonstrate that timed EGFR-TKI administration (≥4 hours before DNA damage induction) potentiates tumor cell killing by prolonging DDR pathway disruption[12, 13]. However, combination regimens are often limited by increased toxicity, complex pharmacokinetics, and challenges in optimizing drug sequencing and dosing. These limitations have spurred growing interest in dual-targeting strategies that simultaneously disrupt EGFR signaling and induce DNA damage. A representative example is ZR2002, a small-molecule agent designed to covalently modify DNA bases via a haloalkyl arm, thereby compromising DNA integrity while concurrently inhibiting EGFR activity. ZR2002 exhibits potent submicromolar anti-proliferative activity even against temozolomide-resistant GSCs [14], highlighting the promise of co-targeting EGFR and DNA.

Phenanthridinone represents an important class of heterocyclic frameworks frequently found in bioactive alkaloids[15] and serves as a core structure with significant potential in antitumor drug discovery[16, 17]. Previously, we synthesized a series of novel phenanthridinone derivatives and identified a crinasiadine-type derivative, ZYH005 (Z5), as the most potent compound, exhibiting even stronger anticancer activity than the well-known phenanthridinone alkaloid narciclasine. In tumor-bearing mouse models, at safe doses, narciclasine significantly inhibited the growth of primary GBM, brain-metastatic melanoma, and lung cancer, but not that of other cancer types[18, 19]. Its relatively high lipophilicity, which facilitates BBB penetration, makes narciclasine a promising candidate for GBM therapy[20]. Another notable phenanthridinone alkaloid, lycorine, has been reported to induce apoptosis in GBM cells through EGFR inhibition and demonstrates efficacy in patient-derived xenograft models[21]. Similarly, narciclasine suppresses gastric cancer cell proliferation via modulation of the AKT/mTOR pathway, which is linked to EGFR signaling[22]. These findings underscore the therapeutic potential of phenanthridinone alkaloids that target EGFR-related mechanisms[22]. Structurally, Z5 possesses fewer hydrophilic hydroxyl groups than narciclasine, suggesting higher theoretical lipophilicity. Consistent with this, we demonstrated that Z5 exhibits superior BBB permeability in mice compared to narciclasine (Supplementary Figure 1A–D). Nevertheless, the anti-GBM efficacy and molecular mechanisms of this novel and more potent phenanthridinone derivatives—particularly their potential interaction with EGFR signaling—remain to be elucidated.

In the present study, we clearly demonstrated that Z5 exerts a significant inhibitory effect on GBM cells and patient-derived glioblastoma stem cells (GSCs) in vitro and in vivo. We confirmed for the first time that Z5 exerts anti-GBM effects through a unique dual-mechanism approach, simultaneously acting as a DNA intercalator and a selective EGFR inhibitor. These findings not only provide a mechanistic foundation for the development of Z5 as a lead compound but also highlight a novel therapeutic hope for GBM, demonstrating compelling potential against this aggressive malignancy and warranting further investigation.

## 2. Materials and Methods

### 2.1 Compounds and cell lines

Z5 was synthesized in our laboratory, and its synthesis process has been described previously[17]. The U87-MG (RRID: CVCL_0022), U251-MG (RRID: CVCL_0021) and HEK293T (RRID: CVCL_0063) cell lines were purchased from Procell Life Science & Technology. GSCs (T3359) were provided by Dr. Shideng Bao and Dr. Jeremy Rich[23]. U87-MG, U251-MG and HEK293T cells were cultured in DMEM (Pricella, Wuhan, China) supplemented with 10% fetal bovine serum (FBS) and 1% penicillin/streptomycin. GSCs were cultured in a serum-free system containing Neurobasal A basal medium supplemented with B-27 supplement minus vitamin A, L-glutamine, sodium pyruvate, 20 ng/ml recombinant epidermal growth factor, and 20 ng/ml basic fibroblast growth factor. The cell culture, cryopreservation, and thawing procedures were all meticulously conducted in strict accordance with standard operating procedures (SOPs). Short tandem repeat (STR) profiling and contamination assessment of the cell lines were performed every six months. The results verified the authenticity of the cell lines and confirmed their contamination-free status.

### 2.2 DNA Microarrays and Gene Set Enrichment Analysis (GSEA)

Total RNA was extracted from U87-MG cells using TRIzol reagent (Cat# 15596-018, Life Technologies, Carlsbad, CA, US). RNA purification was performed using the NucleoSpin RNA Clean-up XS kit (Cat#740903, MN, Germany) and the RNase-Free DNase Set (Cat#79254, QIAGEN, GmBH, Germany). According to the Affymetrix technical manual, the double-stranded cDNA was hybridized with biotin-labeled fragmented cRNA. Microarray experiments were performed using Affymetrix chips by Shanghai Biotechnology Corporation. The raw data were normalized via the expression console algorithm in Command Console Software 4.0 (Affymetrix, Santa Clara, CA, US). Differential expression analysis was conducted via the affy package in R. Genes with an absolute log2-fold change >1 and a P value <0.05 were considered differentially expressed. The microarray data for U87-MG cells have been submitted to the NGDC/OMIX under accession number OMIX008765. Significantly differentially expressed genes (|Log2FC| ≥ 1, FDR ≤ 0.05) are listed in **Supplementary Table 1**. All genes identified through DNA microarray analysis were imported into GSEA software for further analysis and visualization.

### 2.4 Molecular Docking of Z5 with the EGFR Kinase Domain

For our molecular docking experiments, we utilized the crystal structure of EGFR (PDB ID 7KXZ)[24]. This structure was preprocessed via the PDB2PQR program[25] to determine the protonation state of titratable residues at pH=7 and subsequently converted to pdbqt format via the prepare_receptor4.py script from AutoDockTools[26]. Next, we prepared Z5 by converting the compounds to pdbqt format (pH 7.0) via Open Babel. Molecular docking scores were used as indicators of the predicted binding affinity.

### 2.5 Limited Proteolysis–Mass Spectrometry (LiP-MS)

LiP-MS was performed as previously published[27]. A detailed description is provided in the **Supplementary Material**.

### 2.6 Tissue Microarray (TMA) and Immunohistochemistry (IHC)

TMAs were obtained from Wuhan Powerful Biology Company. The use of human tissues in this study was approved by the Ethics Committee of Tongji Hospital of Tongji Medical College of Huazhong University of Science and Technology (Serial number: 2021-lEC-A244). TMA sections were deparaffinized and subjected to antigen retrieval prior to immunostaining. WEE1 antibody (Cell Signaling Technology, 1:200 dilution) was used to detect the protein in the GBM tissue samples.

IHC for in situ transplanted mouse brain tissues was carried out via the same protocol as described above. WEE1 antibody (Cell Signaling Technology, 1:200 dilution) and EGFR antibody (Proteintech, 1:100 dilution) were used to detect the protein.

### 2.7 Orthotopic transplantation mouse model

Experimental BALB/c nude mice were purchased from Beijing Vital River Laboratory Animal Technology Co., Ltd. (Beijing, China) and kept at the Experimental Animal Center of Tongji Medical College, Huazhong University of Science and Technology, Wuhan, China. All the mice were anesthetized with phenobarbital sodium before surgery. One week after GSCs (2×10^4^ cells/mouse) were implanted in the right frontal lobe of 4-week-old mice, 15 mice were randomly divided into three groups: the untreated (NT), Z5 low-dose (10 mg/kg) and Z5 high-dose (20 mg/kg) groups. The mice received either solvent (5% DMSO, 20% castor oil, and 75% saline) or varying doses of the drug by oral gavage daily for 7 weeks. When the NT group showed the first manifestation of obvious neurological signs, one mouse from each of the remaining two groups was evaluated for drug efficacy. The remaining 12 mice were used for survival studies.

### 2.8 Statistical analysis

Statistical analysis was performed via GraphPad Prism 9.0 software. The data are expressed as the means ± SDs. We used unpaired t tests for statistical analysis between 2 groups and one-way ANOVA for multiple comparisons. All the experiments were repeated more than 3 times. We considered a p value less than 0.05 to indicate statistical significance.

## 3. Results

### 3.1 Z5 suppresses proliferation and inhibits EGFRDmediated signaling in GBM cell lines

Using two representative GBM cell lines, U87-MG and U251-MG, we demonstrated that Z5 potently inhibits GBM cell proliferation in a time- and concentration-dependent manner. After 72 hours of treatment, the IC□□ values were 0.114 ± 0.016 μM for U87-MG and 0.147 ± 0.049 μM for U251-MG cells (Figure 1A). Notably, treated with 0.2 μM Z5 for 48 h resulted in approximately 50% inhibition of cell proliferation in both cell lines, whereas the IC_50_ value of lycorine in U251-MG cells was 10 μM under the same conditions[21], indicating that Z5 exhibits superior antiproliferative activity to lycorine in GBM cells.

**Figure 1.**
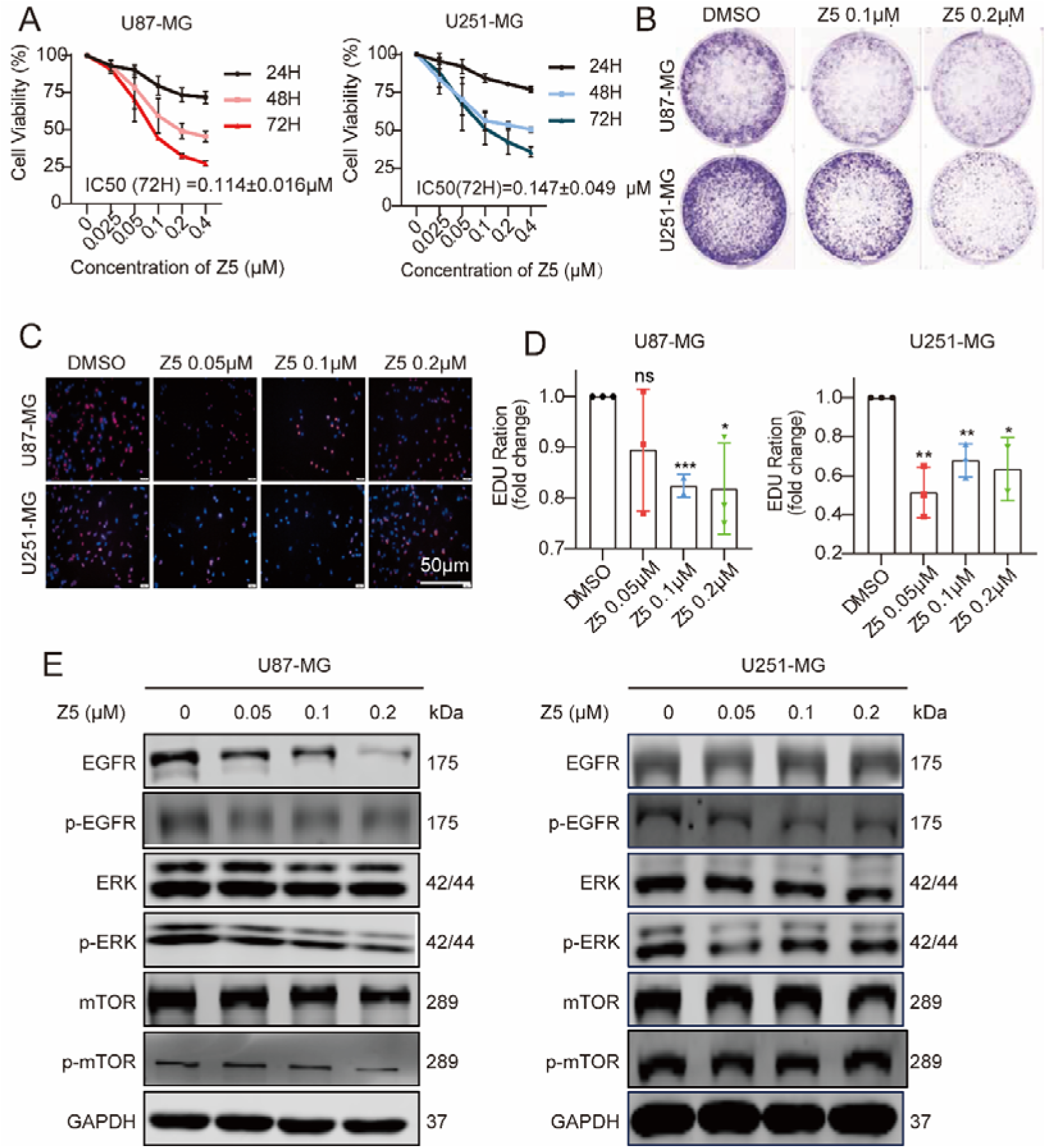
Z5 suppresses proliferation and blocks EGFR-mediated signaling in GBM cell lines. (A) Cell viability of GBM cell lines was assessed by CCK-8 assays after treatment with Z5 for 24, 48, and 72 hours. (B) The inhibitory effect of Z5 was evaluated using the colony formation assay. (C) Fluorescence images showing EdU incorporation in GBM cells under various treatment conditions (red: EdU staining; blue: Hoechst staining). (D) Quantification of EdU-positive cells in each group. The data are presented as the means ± SDs; n=3, compared with the DMSO group. (E) The expression levels of EGFR, p-EGFR, ERK, p-ERK, mTOR and p-mTOR were analyzed by immunoblotting after Z5 treatment.

Furthermore, Z5 significantly suppressed colony formation in both cell lines (Figure 1B). The EdU incorporation assay further demonstrated marked inhibition of DNA synthesis following Z5 treatment, which is key characteristics associated with suppressed cell proliferation (Figure 1C–D). Consistent with its antiproliferative effects and similar to the activity of lycorine[21], treatment with Z5 also significantly suppressed the EGFR signaling pathway, as demonstrated by reduced phosphorylation levels of key biomarkers including EGFR (p-EGFR), ERK (p-ERK), and mTOR (p-mTOR) (Figure 1E).

### 3.2 Z5 suppresses GBM partly through selective targeting of EGFR

To determine whether Z5 directly targets EGFR, we performed a series of biochemical assays. Cellular thermal shift assay (CETSA) demonstrated that Z5 enhanced the thermal stability of EGFR in both U87-MG and U251-MG cell lines (Figure 2A), whereas drug affinity responsive target stability (DARTS) assay also revealed that Z5 protected EGFR from proteolytic degradation (Figure 2B). Most conclusively, surface plasmon resonance (SPR) experiments confirmed direct binding between Z5 and the EGFR kinase domain, with a dissociation constant (KD) of 19.6 μM (Figure 2C).

**Figure 2.**
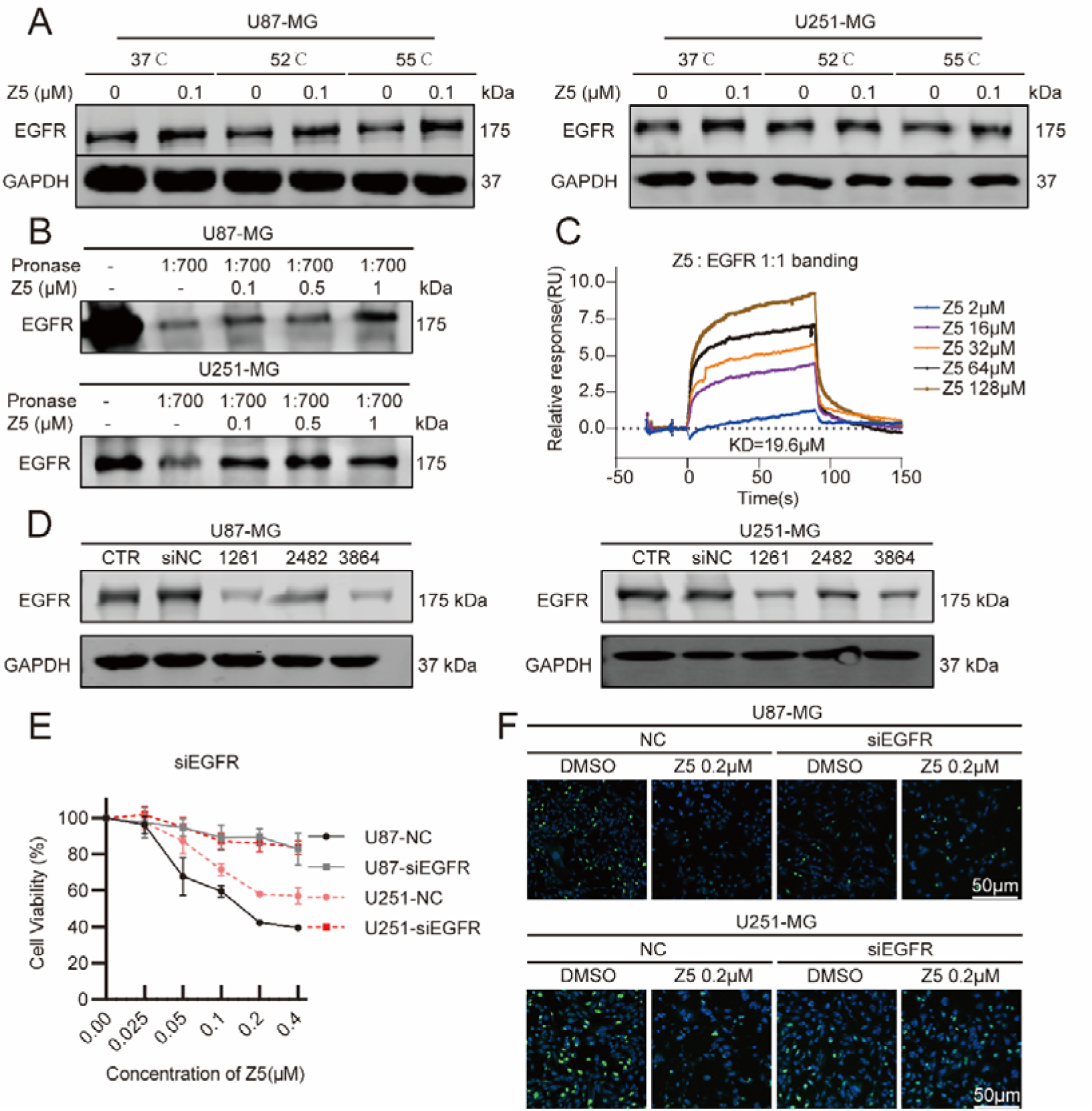
Z5 suppresses GBM partly through selective targeting of EGFR. (A) CETSA assay was performed to assess target engagement of Z5 (0.1 mM) with EGFR in cells. EGFR expression was detected by immunoblotting. (B) DARTS assay was used to evaluate the direct binding of Z5 to EGFR at varying concentrations. EGFR expression was detected by immunoblotting. (C) The direct binding affinity between Z5 and the recombinant EGFR kinase domain protein was analyzed by SPR. (D) EGFR silencing efficiency was evaluated by immunoblot analysis after transfection with EGFR siRNA. (E) Growth curves of NC and si-EGFR-1261 GBM cell lines were measured following treatment with Z5 for 48 h. (F) The anti-proliferative effect of Z5 in NC and si-EGFR-1261 GBM cells was assessed by EdU assay.

To confirm the interaction between Z5 and EGFR, we examined how EGFR silencing influences the antiproliferative effects induced by Z5. The knockdown efficiency of EGFR-targeting siRNAs was first evaluated in GBM cell lines. Based on its superior performance, siEGFR-1261 was selected for further experiments (Figure 2D). Our results demonstrate that EGFR knockdown attenuated—but did not abolish—the anti-proliferative effect of Z5, as evidenced by CCK-8 and EdU assays (Figure 2E–F). Together, these findings establish that Z5 targets EGFR and that its anti-GBM efficacy is largely mediated through this interaction.

### 3.3 Z5 binds to the E762 residue of EGFR

Having established that Z5 directly targets EGFR, we further investigated the structural basis of this interaction. Molecular docking simulations were performed targeting the ATP-binding pocket of the EGFR kinase domain (PDB: 7KXZ). Detailed analysis of the binding mode revealed key molecular interactions between Z5 and EGFR: (1) extensive hydrophobic contacts with residues V702, L820, and L694, stabilizing its position within the ATP-binding pocket; and (2) two critical hydrogen bonds formed with K721 and T766, which likely contribute to its high binding affinity (Figure 3A).

**Figure 3.**
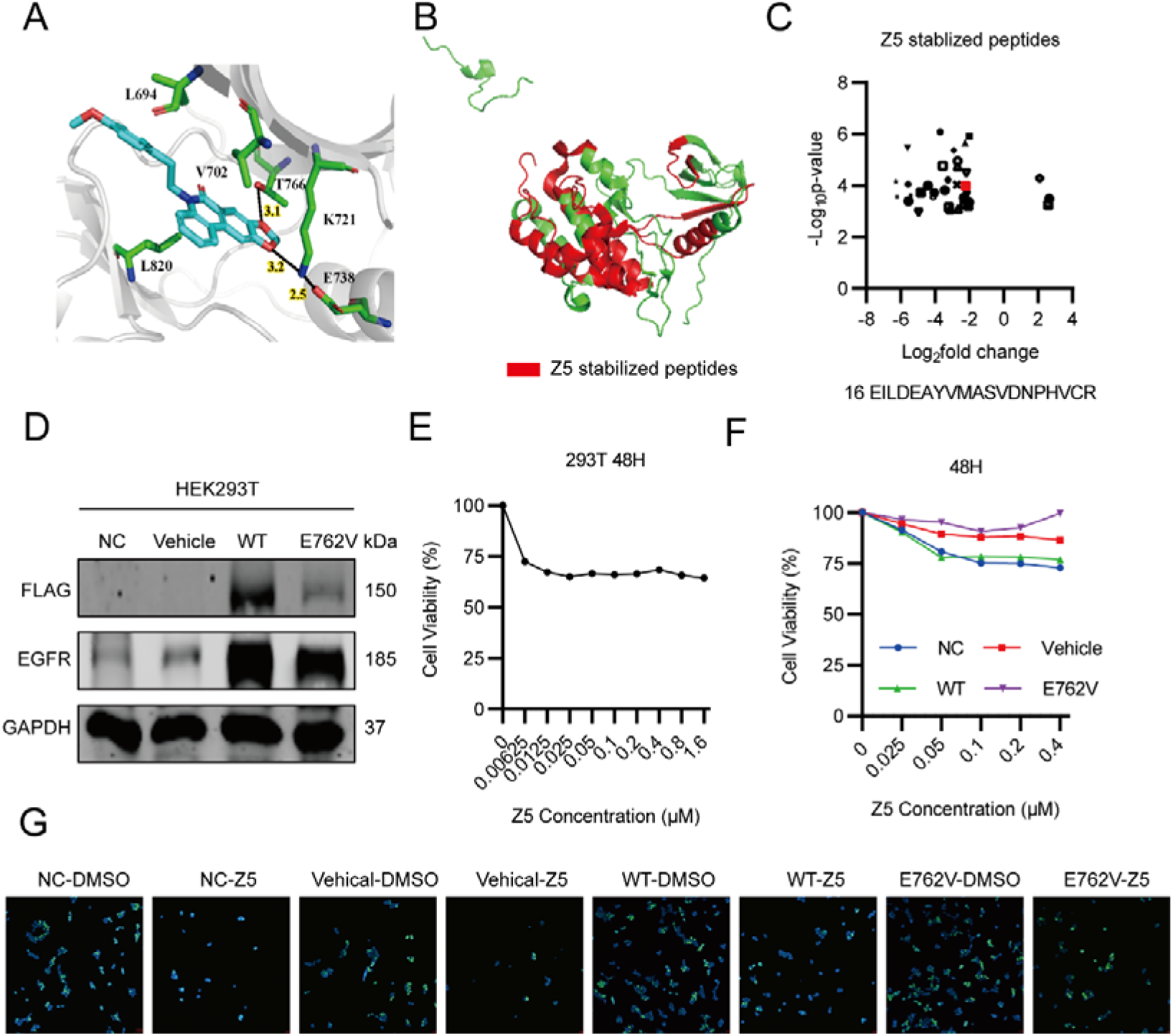
Z5 binds to the E762 residue of EGFR. (A) Predicted binding mode of Z5 within the EGFR kinase domain. (B) Red regions represent peptides with increased proteolytic susceptibility upon Z5 treatment. The selection criteria were as follows: log2(fold change) > 2 and −log10(P value) > 2. (C) Scatter plot displaying changes in peptide abundance from Lip-MS analysis. Each point represents a peptide, with fold-change differences shown between Z5-treated and control samples. (D) Expression of Flag-tagged EGFR and total EGFR in HEK293T cells was analyzed by immunoblotting after transfection with different plasmids. (E) Growth curves of HEK293T cells following treatment with Z5 for 48 h. (F) Growth curves of HEK293T cells transfected with empty vector (vehicle), wild-type (WT) EGFR, or the E762V mutant EGFR, following treatment with Z5 for 48 h. (G) Fluorescence images of proliferating GBM cell lines under various treatment conditions (green: EdU staining; blue: Hoechst staining).

To experimentally validate these computational predictions, we employed limited proteolysis□mass spectrometry (LiP□MS), a label-free proteomics approach, to map potential binding sites of Z5 within the EGFR kinase domain. The LiP-MS analysis identified several peptide segments that are likely involved in Z5 binding (Figure 3B-C). Although the residues K721 and T766—previously implicated in hydrogen bonding by docking studies—were not directly detected by LiP–MS, we observed that K721 forms a hydrogen bond with E738. This interaction may stabilize K721 and thereby indirectly support its hydrogen bonding with Z5. Based on these integrated findings, we propose E738 (corresponding to E762 in UniProt entry P00533-1) as a critical binding residue for Z5. To functionally validate the significance of E762, we established an ectopic expression system in HEK293T cells transfected with empty vector, wild-type (WT) EGFR, or the E762V mutant EGFR (Figure 3D). Dose□response experiments using CCK-8 assay revealed that the antiproliferative effect of Z5 plateaued at concentrations above 0.1 μM, with residual cell viability stabilizing around 75% (Figure 3E). Based on these observations, we selected 0.2 μM Z5 for subsequent experiments to ensure detectable antiproliferative effects while avoiding complete cytotoxicity. Functional validation using both CCK-8 and EdU assays demonstrated that the E762V mutation significantly compromised the antiproliferative effect of Z5 (Figure 3F-G). Compared with cells expressing WT-EGFR, those carrying the E762V mutation exhibited markedly higher cell viability and enhanced proliferative capacity after Z5 treatment. These results confirm that E762 serves as a critical binding residue for Z5 on EGFR and is essential for mediating its antiproliferative activity.

### 3.4 Z5 induces DNA damage through DNA intercalation and EGFR inhibition

To gain deeper insights into the molecular mechanisms underlying Z5-mediated EGFR inhibition, we conducted a comprehensive microarray analysis in U87-MG cells treated with Z5. The analysis identified a total of 1,970 differentially expressed genes (DEGs), among which 188 were upregulated and 1,782 were downregulated (Supplementary Figure 2A). Notably, PRIM1, PRKDC, and MKI67 exhibited the most pronounced downregulation (Supplementary Figures 2B–C).

We next interrogated the microarray data using the Connectivity Map (CMap) database. Querying CMap with a signature comprising the top 66 upregulated and top 150 downregulated genes from our microarray data yielded positive connectivity scores for multiple compounds. We categorized the data by cell type and selected only samples treated with inhibitor perturbations to identify the top ten most effective inhibitors in each cell type. The results showed that EGFR inhibitors appeared most frequently, indicating that the transcriptional impact of Z5 treatment resembles that of multiple known EGFR inhibitors (Figure 4A). This conclusion was further supported by GSEA analysis: Z5 significantly suppressed the activation of ERBB signaling, PI3K/AKT/mTOR pathway, and mTORC1 activity in cells (Figure 4B)—all of which are established pharmacological effects of EGFR inhibition. Together, these data demonstrate that the effect of Z5 on GBM cells is strongly associated with EGFR suppression, further confirming that Z5 acts as a targeted EGFR inhibitor.

**Figure 4.**
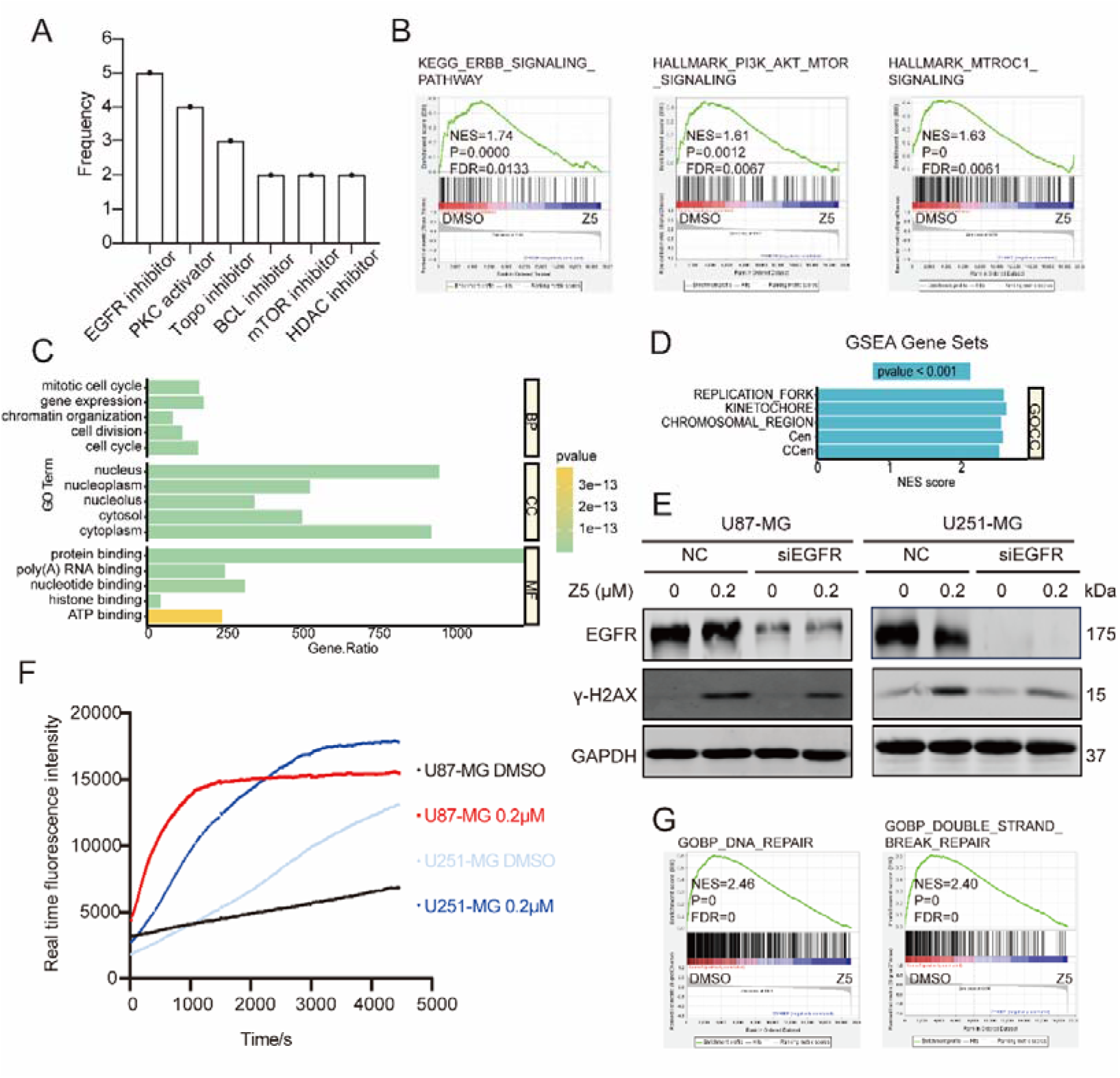
Z5 induces DNA damage through DNA intercalation and EGFR inhibition. (A) From CMAP data, the top 10 compounds positively correlated with microarray profiles were selected per cell line. Compound descriptions were statistically analyzed and ranked by term frequency. (B) GSEA of the ERBB signaling pathway, PI3K/AKT/mTOR signaling and mTORC1 signaling. (C) Top 5 enriched GO terms among downregulated genes in U87-MG cells. (D) GSEA analysis of all genes identified by the U87-MG microarray. Cen: chromosome centromeric region; CCen: condensed chromosome centromeric region. (E) EGFR and γ-H2AX expression in NC and si-EGFR-1261 GBM cells was analyzed by immunoblotting after 48 h Z5 treatment. (F) Increased APE1-accessible DNA damage in Z5 treated cells. (G) GSEA of the DNA repair and double strand break repair.

Cellular Component (CC) enrichment analysis from GO and GSEA further revealed significant enrichment of dysregulated genes involved in nuclear compartments, replication forks, and chromosomal regions following Z5 treatment (Figure 4C–D), consistent with its DNA intercalating properties. In line with these findings, Z5 treatment induced significant DNA damage in GBM cells, as evidenced by increased APE1-mediated cleavage events—detected using a dual-amplification fluorescent biosensor[28]—and elevated γH2AX foci indicating double-strand breaks (Figure 4E–F). These findings suggest that the nucleus serves as a primary site for Z5’s mechanism of action.

It is well-established that nuclear EGFR plays a critical role in the DDR, and that EGFR inhibition impairs DDR, resulting in accumulation of DNA damage. In line with this, GSEA showed significant downregulation of DNA repair pathways after Z5 treatment (Figure 4G, Supplementary Figure 2D), confirming that Z5 acts as an EGFR inhibitor to suppress the DDR. To further examine whether EGFR contributes to Z5-induced DNA damage, we knocked down EGFR using siEGFR-1261. First, we found that EGFR knockdown alone—despite its potential to suppress DDR—did not induce DNA damage in the absence of Z5 treatment. Second, EGFR knockdown reduced Z5-induced DNA damage compared to wild-type cells (Figure 4E), indicating that EGFR plays a functional role in Z5-mediated DNA damage. Specifically, EGFR inhibition by Z5 acts synergistically to amplify DNA damage initiated through its primary mechanism of DNA intercalation. Together, these results demonstrate that Z5 induces DNA damage via a dual mechanism involving both DNA intercalation and EGFR inhibition.

### 3.5 Z5 induces G2/M arrest and WEE1-mediated mitosis catastrophe through EGFR inhibition

Following DNA damage, cells typically initiate cell cycle arrest to allow time for repair. Consistent with this mechanism, KEGG and GO analyses indicated that Z5 predominantly affects pathways related to cell cycle regulation and mitotic progression (Figure 5A). Subsequent GSEA using the GO-BP, HALLMARK, and KEGG databases further underscored significant enrichment of terms associated with G2/M checkpoint signaling, G2/M phase transition, cell cycle control, and mitotic spindle assembly (Figure 5B, Supplementary Figure 2E). These findings suggest that Z5 disrupts the tightly regulated G2/M transition process. Consistent with these findings, we observed a robust arrest in the G2/M phase by flow cytometry (Figure 5C). Notably, Z5 treatment markedly elevated the populations of PHH3-positive and polyploid cells (Figure 5D), a hallmark of mitotic catastrophe that is consistent with checkpoint failure. In addition, we found that EGFR knockdown completely abolished Z5-induced G2/M arrest, demonstrating that Z5-mediated cell cycle blockade is strictly dependent on EGFR inhibition (Figure 5E). To elucidate how Z5 disrupts the G2/M checkpoint through EGFR, we analyzed DEGs from microarray data. Among key regulators of the G2/M checkpoint, WEE1 showed the most pronounced downregulation (67%), while CDC2 (also known as CDK1)—which is phosphorylated by WEE1 to regulate mitotic entry—was also substantially reduced (55%) (Supplementary Table 1), suggesting transcriptional disruption of the WEE1-mediated checkpoint by Z5. Corroborating these findings, immunoblot analysis showed decreased phosphorylation of WEE1 and CDC2, along with upregulated PHH3 expression following Z5 treatment (Figure 5F). Furthermore, immunofluorescence (IF) assays unveiled hallmark morphological features of mitotic catastrophe, such as chromosomal mis-segregation, multipolar spindle formation, and micronucleation (Figure 5G). Collectively, these results demonstrated that Z5 induces G2/M arrest and mitotic catastrophe through EGFR-mediated inhibition of the WEE1 checkpoint.

**Figure 5.**
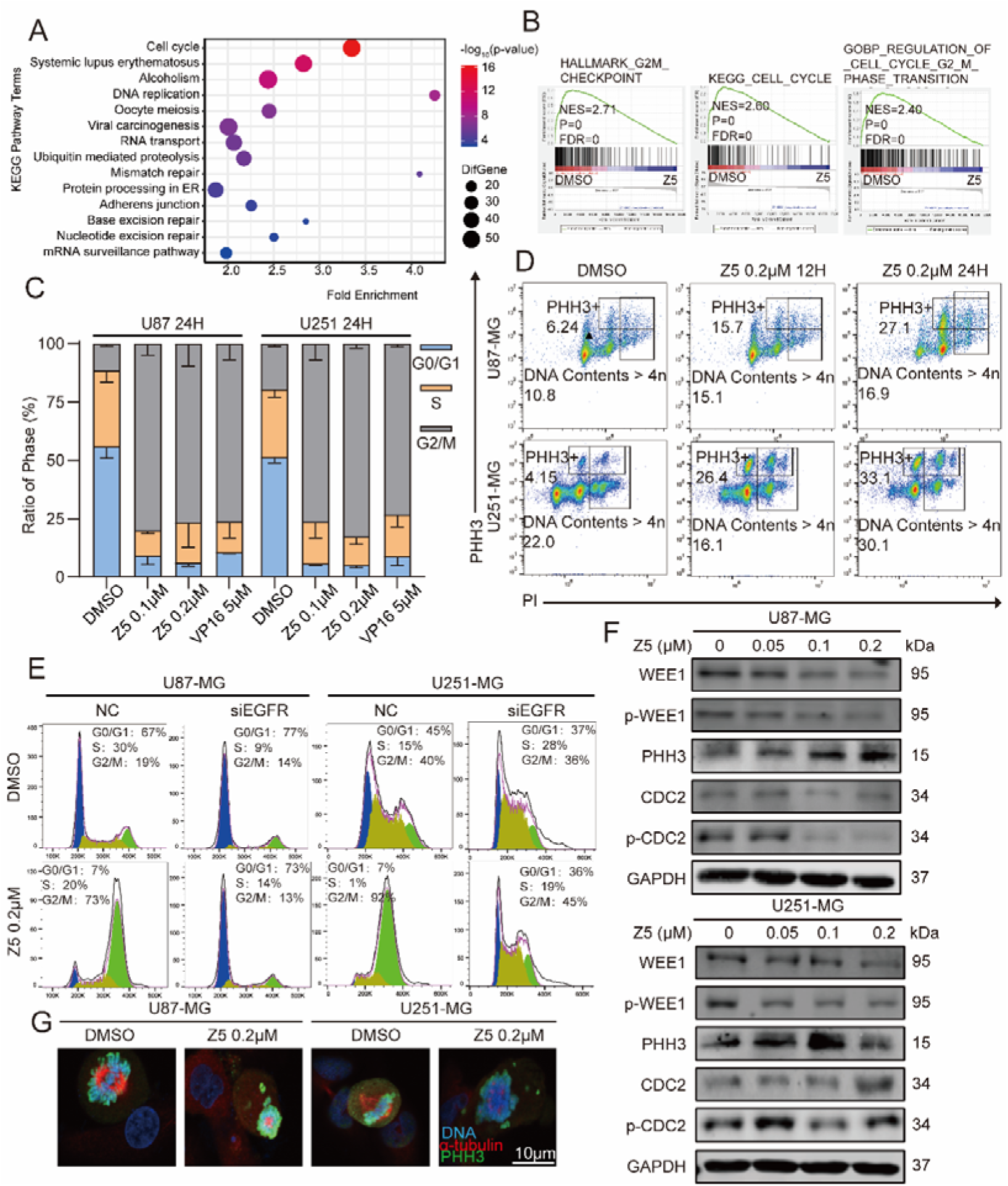
Z5 induces G2/M arrest and mitotic catastrophe in GBM cells. (A) KEGG pathway enrichment analysis of microarray data from GBM cells. (B) GSEA of the G2M checkpoint, cell cycle and cell cycle G2/M phase transition pathways. (C) Cell cycle distribution in GBM cell lines was analyzed by flow cytometry after 24 h Z5 treatment. (D) Representative flow cytometric analysis of PHH3+ and DNA contents > 4n cells. (E) Cell cycle profiles of NC and si-EGFR GBM cells were assessed by flow cytometry after 24 h Z5 treatment. (F) Expression of WEE1, p-WEE1, PHH3, CDC2, and p-CDC2 was analyzed by immunoblotting after 48 h treatment with increasing concentrations of Z5. (G) IF staining was performed with anti-tubulin (red) and anti-PHH3 (green) antibodies in U87-MG and U251-MG cells treated with DMSO or Z5.

### 3.6 Z5 targets nuclear EGFR to disrupt the EGFR-WEE1 axis

To investigate whether the anti-GBM efficacy of Z5 involves functional crosstalk between EGFR and WEE1, we first analyzed their correlation in glioma transcriptomes. Data from the Chinese Glioma Genome Atlas (CGGA) revealed a significant positive correlation between EGFR and WEE1 mRNA expression (Pearson’s R = 0.38; Figure 6A). To determine whether this co-expression reflects direct molecular interaction, we performed SPR analysis, which demonstrated high-affinity binding between the kinase domains of EGFR and WEE1 with an equilibrium dissociation constant (KD) of 17.3 nM (Figure 6B). This interaction was further confirmed by co-immunoprecipitation (Co-IP) in GBM cells. Importantly, Z5 treatment significantly disrupted the EGFR–WEE1 complex, as shown by reduced Co-IP of both proteins (Figure 6C; Supplementary Figure 3A).

**Figure 6.**
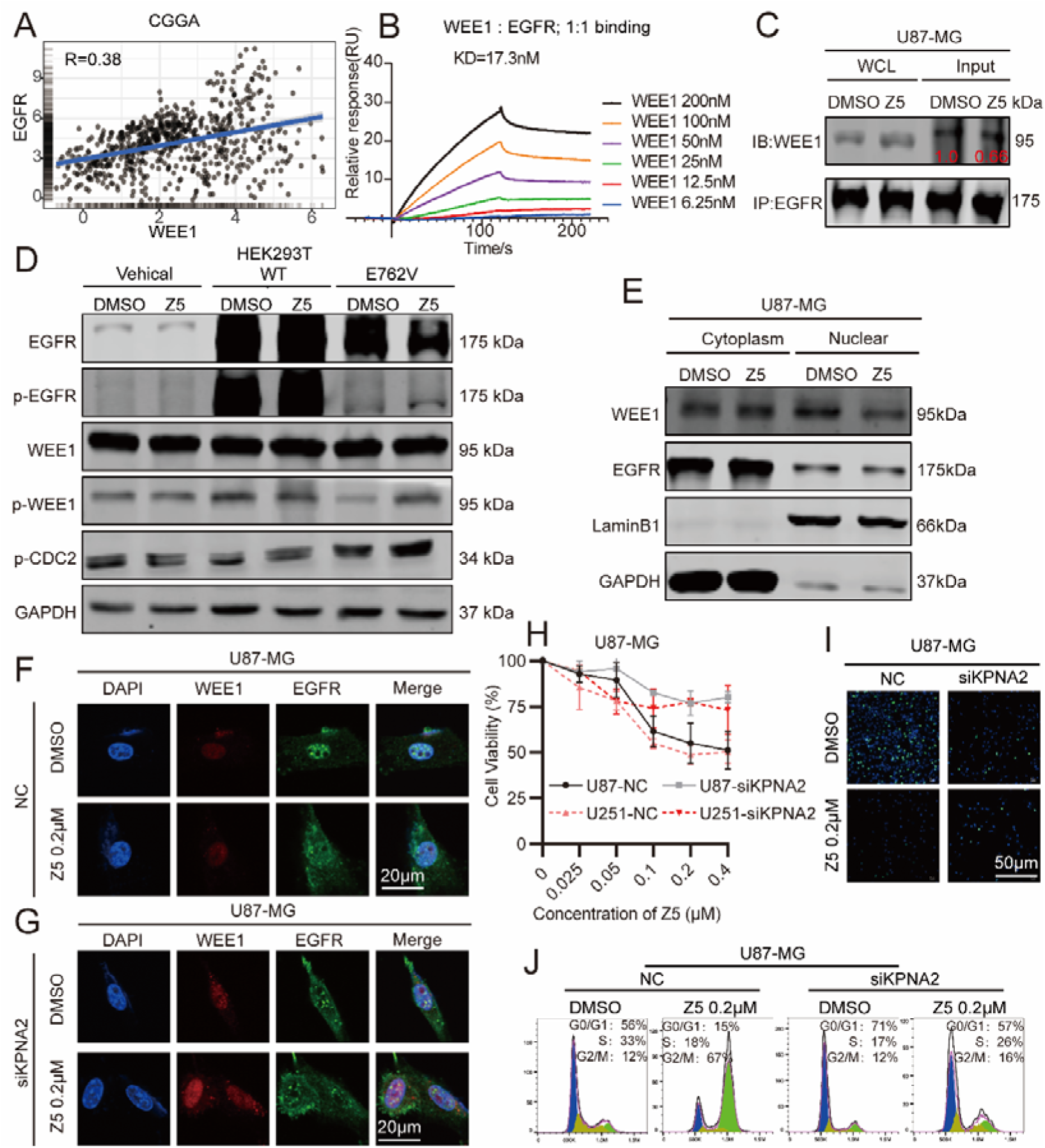
Z5 disrupt the nuclear EGFR-WEE1 axis. (A) Correlation between EGFR and WEE1 expression in GBM patients. (B) SPR analysis of the interaction between the kinase domains of EGFR and WEE1. (C) Co-IP was performed to assess EGFR–WEE1 interaction in U87-MG cells after 15 min Z5 treatment. (D) Western blot analysis of WEE1, p-WEE1, p-CDC2, EGFR, and p-EGFR in HEK293T cells transfected with empty vector, WT-EGFR, or E762V-EGFR. (E) Subcellular localization of EGFR and WEE1 in cytoplasm and nucleus was analyzed in U87-MG cells after 15 min Z5 treatment. (F-G) IF staining of EGFR and WEE1 in NC and siKPNA2-1400 U87-MG cells. (H) Growth curves of NC and siKPNA2-1400 GBM cells after 48 h Z5 treatment. (I) Anti-proliferative effect of Z5 was assessed by EdU staining in NC and siKPNA2-1400 U87-MG cells. (J) Cell cycle distribution was analyzed by flow cytometry in NC and siKPNA2-1400 U87-MG cells after 24 h Z5 treatment.

Next, we performed genetic and pharmacological perturbations in U87-MG cells to dissect the functional relationship between EGFR and WEE1. Knockdown of WEE1 did not affect EGFR expression or phosphorylation. In contrast, EGFR knockdown significantly reduced both total WEE1 and phospho-WEE1 levels, establishing EGFR as an upstream regulator of WEE1 (Supplementary Figure 3B). Consistent with these findings, the WEE1 inhibitor MK1775 had minimal impact on EGFR signaling, whereas the EGFR inhibitor AZD9291 strongly suppressed both p-WEE1 and p-CDC2. These results confirm a unidirectional regulatory axis from EGFR to WEE1 (Supplementary Figure 3C). Building on this established EGFR to WEE1 signaling relationship, we further investigated whether Z5 exerts its effects through direct interaction with EGFR. In 293T cells expressing the E762V mutant EGFR, Z5-induced downregulation of p-WEE1 and p-CDC2 was markedly attenuated compared to wild-type controls (Figure 6D). This indicates that Z5 targets EGFR via the E762 residue to inhibit its activity and disrupt the EGFR–WEE1 regulatory axis.

Since EGFR undergoes nucleocytoplasmic shuttling and WEE1 is predominantly nuclear, we hypothesized that their interaction is nuclear-specific. To test this, we assessed their co-localization. In control (DMSO-treated) cells, both IF and cellular fractionation confirmed substantial nuclear co-localization of EGFR and WEE1. Z5 treatment, however, disrupted this co-localization, resulting in the dissociation of both proteins and their accumulation in the cytoplasm (Figure 6E–F; Supplementary Figure 3D).

We next investigated whether the nuclear import of EGFR is essential for the anti-GBM effects of Z5. Given that KPNA2 and KPNB1 facilitates EGFR nuclear translocation [29], we knocked KPNA2 down using a specific siRNA (siKPNA2-1400) and confirmed a subsequent reduction in nuclear EGFR levels (Figure 6G; Supplementary Figure 3E). This intervention significantly attenuated the ability of Z5 to inhibit cell proliferation (as assessed by CCK-8 and EdU assays; Figure 6H-I, Supplementary Figure 3F) and to induce G2/M phase arrest (Figure 6J; Supplementary Figure 3G). The critical role of nuclear import was further emphasized when pharmacological inhibition of KPNB1 with importazole synergized with Z5 to further diminish its efficacy (Supplementary Figure 3F). Importantly, the partial anti-proliferative effect that remained after KPNA2 knockdown suggests that Z5’s mechanism of action also involves pathways dependent on cytoplasmic and membrane-localized EGFR. This finding is also in line with our previous detection of Z5’s inhibition of the EGFR-downstream mTOR and ERK pathways.

Given the limited exploration of the EGFR-WEE1 interaction in GBM, especially within clinical samples, we sought to address this gap by conducting an in-depth analysis of transcriptomic data from GBM patient cohorts. This analysis aimed to evaluate the clinical relevance of this axis and to assess the translational potential of Z5. In the CGGA database, WEE1 expression was significantly elevated in GBM compared to other glioma subtypes (Supplementary Figure 4A), with classical and mesenchymal subtypes showing higher levels than the proneural subtype (Supplementary Figure 4B). IDH-wildtype GBM exhibited stronger WEE1 expression relative to IDH-mutant tumors (Supplementary Figure 4C), consistent with a more aggressive phenotype. Survival analyses using TCGA and CGGA datasets revealed that high WEE1 expression was associated with poorer overall survival in GBM patients (Supplementary Figure 4D–F). At the protein level, immunohistochemical staining of a tissue microarray containing 64 primary GBM samples (grades 0–IV) showed a positive correlation between WEE1 intensity and tumor grade (Supplementary Figure 4G–H). IF further confirmed colocalization of EGFR and WEE1 in GBM tissues (Supplementary Figure 4I), supporting their functional interaction in vivo.

To summarize, we established that EGFR acts as an upstream regulator of WEE1 and revealed their specific nuclear interaction as a critical vulnerability. Z5 binds to EGFR, disrupts the nuclear EGFR-WEE1 complex, and consequently inhibits downstream signaling, leading to cell cycle arrest and suppressed tumor growth. The clinical relevance of this axis, supported by its association with aggressive tumor subtypes and poorer patient survival, highlighting the central role of the EGFR–WEE1 axis in GBM pathogenesis and the translational potential of Z5.

### 3.7 Z5 suppresses GSCs tumorigenesis and inhibits EGFR-related signaling in vivo

GSCs represent a distinct subpopulation within GBM, defined by their stem-like properties and enhanced DNA repair capacity [30, 31]. Given their critical role in treatment resistance and tumor recurrence, we chose this most challenging and representative model to further assess Z5’s anti-GBM effects. Accordingly, we performed tumor sphere formation assay, which revealed that Z5 markedly induced fragmentation of tumor spheres derived from the T3359 GSCs (Figure 7A–B). Immunoblotting further confirmed that Z5 significantly suppressed the EGFR–WEE1axis in these cells (Figure 7C).

**Figure 7.**
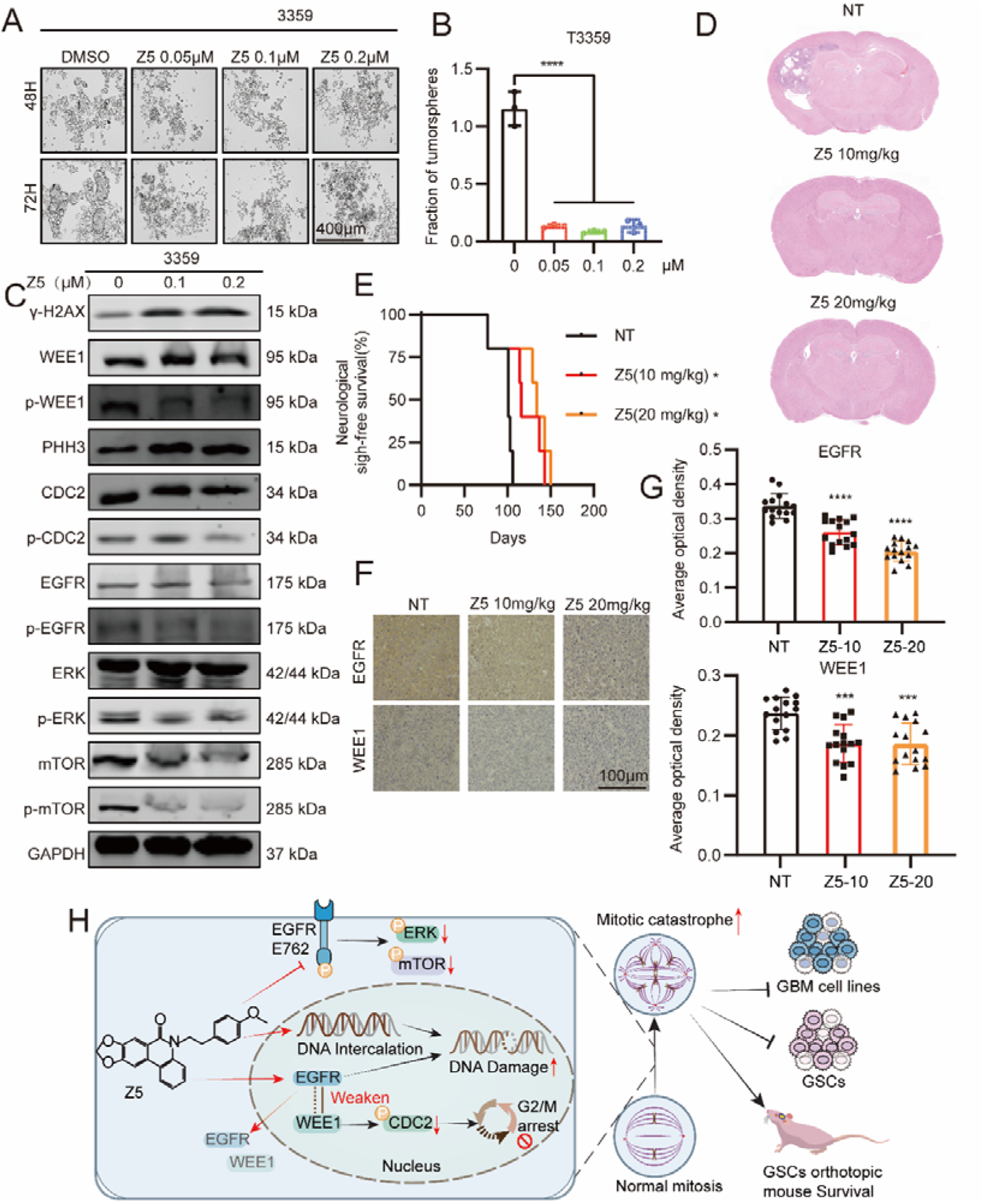
Z5 suppresses GSCs tumorigenesis and inhibits EGFR-related signaling in vivo. (A) Representative tumor sphere formation assay in T3359 cells treated with different concentrations of Z5 for 48 h. Scale bar = 400 μm. (B) Quantitative analysis of tumor sphere fragmentation from (A). The data are presented as the means ± SDs, n=3, ****p < 0.0001, compared with the “0” group. (C) Expression of γ-H2AX, WEE1, p-WEE1, PHH3, CDC2, p-CDC2, EGFR, p-EGFR, ERK, p-ERK, mTOR, and p-mTOR in T3359 cells with DMSO or Z5 treatment. (D) H&E staining of intracranial xenografts to assess tumor formation. (E) Survival analysis of mice orthotopically implanted with GSCs. The data are presented as the means ± SDs, n=4, *p < 0.05, compared with the NT group. (F) Immunohistochemical analysis of EGFR and WEE1 expression in orthotopic tumor tissues. (G) Quantification of EGFR and WEE1 levels by average optical density from five fields per mouse. The data are presented as the means ± SDs, n=3, ****p < 0.0001, compared with the NT group. (H) Schematic diagram illustrating the mechanism of Z5.

For in vivo validation, we established an orthotopic xenograft model. Upon terminal symptom onset in the first untreated (NT) mouse, one mouse per treatment group was euthanized for histological analysis. HE staining showed that Z5 markedly suppressed intracranial GSCs tumor growth (Figure 7D). Z5 also prolonged survival in a dose-dependent manner. Mice in the NT group survived for a median of 102.75 days, whereas those treated with 10 mg/kg and 20 mg/kg Z5 exhibited median survivals of 127.5 and 139 days, corresponding to 24.1% and 35.3% increases, respectively (Figure 7E). IHC staining further demonstrated reduced EGFR and WEE1 expression in Z5-treated tumors (Figure 7F–G). Notably, after 8 weeks of treatment, H&E staining of heart, liver, spleen, lung, and kidney tissues revealed no toxicity (Supplementary Figures S5A–E). These findings establish Z5 as a potent inhibitor of GSCs growth in both in vitro and in vivo settings, accompanied by effective suppression of EGFR-WEE1 axis and a favorable safety profile.

In conclusion, Z5 exerts potent anti-GBM efficacy via a dual mechanism: DNA intercalation that induces damage, and targeted inhibition of EGFR at E762. The DNA damage response is amplified by EGFR inhibition, which concurrently disrupts the EGFR-WEE1 interaction. This disruption compromises WEE1-mediated regulation of its downstream effector CDC2, leading to reduced p-CDC2 levels, G2/M checkpoint failure, mitotic aberrations, and ultimately, mitotic catastrophe. Furthermore, Z5 inhibits the mTOR and ERK pathways, confirming the suppression of plasma membrane-localized EGFR signaling as a key contributor to its efficacy. Collectively, based on these mechanisms, Z5 demonstrates potent anti-tumor activity across diverse preclinical GBM models, including established cell lines, patient-derived GSCs, and orthotopic xenografts, underscoring its considerable therapeutic potential (Figure 7H).

## 4. Discussion

Since TMZ’s approval in 2005, despite extensive research over the past 2 decades, no transformative therapies have emerged. Currently, the top-priority task in this field is to spare no effort in the search for new molecules and methods that are effective against GBM. Addressing this critical gap, our study successfully identified Z5 as a novel, dual-functional EGFR inhibitor that not only demonstrates potent anti-tumor efficacy across a spectrum of preclinical GBM models but also exhibits a favorable safety profile, distinguishing it from many existing therapeutic candidates. More importantly, we have systematically elucidated its unique mechanism of action, revealing that Z5 achieves its robust effects by simultaneously inducing DNA damage through intercalation and precisely targeting the EGFR signaling axis at the E762 residue, thereby disrupting the critical EGFR-WEE1 regulatory circuit and downstream survival pathways. This comprehensive investigation positions Z5 as a highly promising candidate worthy of further clinical development, offering a new strategic approach to combat this devastating disease.

We have previously established Z5 as a DNA intercalator [17]. Z5 exhibits moderate intercalation strength compared to classical intercalators such as YOYO-1 and ethidium bromide. This property may correlate with reduced carcinogenic and mutagenic potential, as supported by in vitro and in vivo toxicity studies in normal cells and mouse models. Single-molecule magnetic tweezer experiments also revealed that Z5 induces only moderate changes in DNA extension and twist under low force conditions, which may account for its potent antitumor activity coupled with low systemic toxicity [17]. These findings were further corroborated in the present study through long-term toxicity assessment over 150 days in an orthotopic GBM model using patient-derived GSCs. Given its high safety profile, further investigation into its full range of biological effects and underlying mechanisms is warranted.

Despite the introduction of novel therapeutic modalities such as tumor-treating fields, molecularly targeted agents, and immunotherapies, the clinical management of GBM remains profoundly challenging. These approaches are consistently hampered by tumor heterogeneity, an immunosuppressive microenvironment, and—critically—the limited penetration of the BBB, which collectively contribute to transient responses and recurrence rates exceeding 90%[32, 33]. In particular, conventional EGFR inhibitors (e.g., gefitinib, osimertinib) exhibit restricted efficacy in GBM, owing not only to compensatory signaling and tumor adaptability but also to their inadequate BBB permeability [34–36]. Z5, as a novel phenanthridine alkaloid derivative, has shown remarkable BBB penetration and potent anti-tumor efficacy, providing further evidence for the ability of this structural class to cross the BBB and exert therapeutic effects in brain diseases. Consistently, natural phenanthridine alkaloids such as narciclasine and lycorine have been confirmed by previous studies to exhibit significant anti-GBM activity, while lycorine, galanthamine, and haemanthamine have also demonstrated neuroprotective effects[37–39]. Of particular note, galanthamine, a representative phenanthridine alkaloid and a natural product from the *Amaryllidaceae* family, has been approved by the FDA for the treatment of Alzheimer’s disease [40]. Its successful clinical application serves to underscore the potential of this chemical family to cross the BBB and treat neurological disorders, thereby offering a compelling rationale and considerable promise for the translational research of Z5.

DNA intercalators reversibly insert between adjacent base pairs of double-stranded DNA, thereby altering DNA conformation, compromising structural integrity, and interfering with DNA-processing proteins [40, 41]. Unlike alkylating agents, which exert anticancer effects through irreversible covalent modification of DNA, intercalators bind via noncovalent interactions—primarily van der Waals forces, hydrogen bonding, hydrophobic effects, and/or charge-transfer interactions—resulting in reversible DNA binding [42]. Consequently, their binding does not necessarily induce immediate cytotoxicity. To elicit cyell-killing effects, intercalators must first form a sufficiently stable complex with a long half-life, capable of obstructing DNA-metabolizing enzymes and impairing essential processes such as replication, transcription, and translation. Classic examples include anthracycline drugs like doxorubicin and daunorubicin, which intercalate into DNA but primarily cause cytotoxicity by inhibiting topoisomerase II, preventing DNA religation and leading to irreversible strand breaks. In this study, we employed multiple experimental approaches to demonstrate that beyond its DNA-intercalating activity, Z5 also functions as a potent EGFR inhibitor. By specifically targeting the E762 residue of EGFR, Z5 effectively suppresses canonical plasma membrane-derived signaling pathways—such as mTOR and ERK—and concurrently impairs EGFR-mediated DNA damage repair, resulting in the accumulation of unresolved DNA lesions. Importantly, this strategy represents a significant advance over monotherapeutic approaches that focus exclusively on nuclear EGFR or individual downstream effectors[43, 44].

WEE1 plays a critical role in maintaining genomic stability by phosphorylating CDK1/2 to enforce the G2/M checkpoint, thereby allowing DNA repair before mitotic entry[45]. Its overexpression is frequently associated with poor prognosis and therapy resistance in multiple cancers, while pharmacological or genetic inhibition of WEE1 can induce mitotic catastrophe in genomically unstable tumor cells[46]. In GBM, where EGFR amplification and replication stress are highly prevalent, WEE1 serves as a key survival factor that helps cancer cells cope with DNA damage and replication pressure[47]. In this study, we systematically delineated, for the first time, the functional interplay between EGFR and WEE1 in GBM. We demonstrated that Z5 disrupts this EGFR–WEE1 axis, thereby compromising the protective checkpoint mechanism and driving GBM cells into mitotic catastrophe. These findings not only reveal a novel mode of action for Z5 but also provide compelling evidence supporting the EGFR–WEE1 axis as a therapeutically targetable vulnerability in GBM. Z5 represents both a mechanistic probe for understanding EGFR–WEE1 biology and a promising therapeutic candidate worthy of further development.

While our study elucidates a novel mechanism of action for Z5, several questions remain unresolved. First, it is still unclear whether the disruptive effects of Z5 on the EGFR–WEE1 interaction are mediated solely through its EGFR-targeting activity, independently of DNA intercalation, or via a combined mechanism. Second, the structural basis of the EGFR–WEE1 interaction—particularly the precise binding interface—requires further elucidation [7, 47, 48]. Third, it remains to be determined whether Z5 inhibits EGFR tyrosine kinase activity and how its efficacy compares with that of existing EGFR inhibitors. These aspects are critical for optimizing the therapeutic application of Z5 and warrant further investigation.

## 5. Conclusions

In conclusion, Z5 demonstrates potent anti-GBM efficacy by concurrently targeting DNA and EGFR, which elicits mitotic catastrophe while maintaining a favorable safety profile. This study establishes Z5 not only as a mechanistic probe for EGFR signaling but also as a brain-penetrant clinical candidate, and reveals disruption of the nuclear EGFR–WEE1 axis as a novel therapeutic vulnerability in GBM. These findings provide a strong foundation for advancing Z5 toward clinical translation and offer a promising new strategy for GBM treatment.

## Data Availability

This study did not generate new reagents or original code. All the data generated or analyzed during this study are included in this article and its supplementary information files. Microarray data can be accessed through the following link: https://doi.org/10.1101/2025.04.14.25325787

https://ngdc.cncb.ac.cn/omix/preview/BlYvj2iS.

## 6 Abbreviations

GBM: Glioblastoma
EGFR: Epidermal growth factor receptor
Z5: ZYH005
GSCs: glioblastoma stem cells
qPCR: Quantitative polymerase chain reaction
γ-H2AX: Phospho-Histone H2A.X
TMZ: Temozolomide
DNA-PKcs: DNA-dependent protein kinase catalytic subunit
NHEJ: nonhomologous end joining
DDR: DNA damage response
TKIs: Tyrosine kinase inhibitors
BBB: Blood–brain barrier
GSCs: Glioblastoma stem cells
FBS: Fetal bovine serum
CTR: Control
SOPs: Standard operating procedures
STR: Short tandem repeat
GSEA: Gene Set Enrichment Analysis
LiP-MS: Limited Proteolysis–Mass Spectrometry
IHC: Immunohistochemistry
TMA: Tissue Microarray
CETSA: Cellular thermal shift assay
DARTS: Drug affinity responsive target stability
SPR: Surface plasmon resonance
GO: Gene Ontology
PHH3: Phospho-Histone H3
H&E: Hematoxylin and eosin
CGGA: Chinese Glioma Genome Atlas
IDH: Isocitrate Dehydrogenase
TCGA: The Cancer Genome Atlas
OS: Overall survival
Co-IP: Co-Immunoprecipitation
NT: Untreated
KEGG: Kyoto Encyclopedia of Genes and Genomes

## 7. Declarations

### 7.1 Ethics approval and consent to participate

GSC cells were obtained from glioma patients who underwent surgery at the Department of Neurosurgery of Tongji Hospital, Tongji Medical College of Huazhong University of Science and Technology. All participants provided written informed consent, and the study was approved by the Ethics Committee of Tongji Hospital of Tongji Medical College of Huazhong University of Science and Technology (Serial number: 2021-lEC-A244). The Animal Experiment Administration Committee of the Huazhong University of Science and Technology (HUST) approved all the animal experiments to ensure ethical and humane treatment of the animals (IACUC number: 3028).

### 7.2 Consent for publication

Not applicable.

### 7.6 Author contributions

Yonghui Zhang, Qingyi Tong, Suojun Zhang and Xingjiang Yu: Funding acquisition, Resources. Jianzheng Huang and Qingyi Tong: Data curation, writing-original draft and writing-review editing. Zijun Zhang, Xiao Yang and Ziming Zhao: Validation. Zengwei Luo, Junjun Liu, Suitian Lai: Visualization. Chao Song and Shouchang Feng: Resources.

## 7.7 Acknowledgments

We thank Dr. Shideng Bao, Dr. Jeremy Rich and Professor Xingjiang Yu for generously providing GSCs (T3359 and D456). We thank the Medical Subcenter of HUST Analytical & Testing Center for acquiring the data and the Experimental Animal Centre of HUST for providing the animal experimentation facilities.

## Supporting Information

### 1. Materials and Methods

#### 1.1 Histology analysis

Brain tissues were fixed with 4% paraformaldehyde for 12 hours and embedded in paraffin. The paraffin-embedded brain specimens were sectioned and stained with H&E, according to the manufacturer’s protocol. Other sections of brain specimens were subjected to immunohistochemistry (IHC), and the samples were incubated with primary antibodies against WEE1 (Cell Signaling Technology) and EGFR (Abclonal) for 30 minutes at room temperature. Subsequently, the slides were incubated with secondary antibodies and hematoxylin.

#### 1.2 Determination of Blood-Brain Barrier (BBB) Penetration

To evaluate the BBB penetration of Z5, twelve C57BL/6 mice were randomly divided into four groups: a vehicle control group and three treatment groups sacrificed at 0.25, 0.5, and 2 h after intraperitoneal administration of Z5. After injection, mice were anesthetized with isoflurane and blood samples (∼0.5 mL) were collected by cardiac puncture into pre-chilled tubes containing EDTA-K□ as an anticoagulant. Samples were kept on wet ice and centrifuged within 1 h of collection (1700 × g, 4□°C, 10 min) to obtain plasma.

Subsequently, mice were transcardially perfused with ice-cold saline (0.9% NaCl) until the liver appeared pale to remove intravascular blood. Brain tissues were then harvested and homogenized in a ratio of 1:3 (w/v; 1 g tissue to 3 mL water). The concentrations of Z5 in plasma and brain homogenate were quantified using liquid chromatography-tandem mass spectrometry (LC-MS/MS).

#### 1.3 Cell viability assay

Cell viability was measured using the CCK-8 assay. Briefly, cells were seeded into 96-well plates at a density of 3,000 cells per well and allowed to adhere overnight before treatment with various concentrations of drugs or vehicle. At 24, 48, or 72 h post-treatment, 10 μL of CCK-8 reagent was added to each well and cells were incubated at 37°C for 4 h. The optical density (OD) was measured at 450 nm using a microplate reader. IC□□ values were determined by nonlinear regression analysis of dose-response curves using SPSS software

For drug combination studies, cells were co-treated with varying concentrations of ZYH005 and importazole for 48 h. Synergy scores were calculated using the SynergyFinder+ web application: <u>SynergyFinder+</u>.

#### 1.4 Clone formation assay

Cells were seeded into 6-well plates at a density of 500 cells per well and allowed to adhere overnight. Cells were then treated with DMSO or Z5 for 12 h, after which the drug-containing medium was replaced with fresh drug-free medium. The medium was changed every three days thereafter. The cells were collected after 14 days of culture.

#### 1.5 EdU staining

EdU cell proliferation staining was performed using an EdU kit (BeyoClick™ EdU Cell Proliferation Kit with Alexa Fluor 488, Beyotime, China). The kit was used according to the manufacturer’s instructions.

#### 1.6 Immunoblotting assay

Immunoblotting was performed as previously published^1^. The membranes were washed with TBST and incubated with the secondary antibodies anti-rabbit IgG (H+L) (CST, #5151, Boston, American) and anti-mouse IgG (H+L) (CST, #5257, Boston, American), followed by imaging using a Li-Cor Odyssey scanner (LI-COR, USA). Image analysis was executed using Image Studio software. Primary antibodies used in the study are listed in **Supplementary Table S2**.

#### 1.7 DNA fragment detection assay

Cells were seeded into 6-well plates and incubated with DMSO or Z5. After 24 hours, the cells were washed once with cold PBS. Genomic DNA was extracted using the Universal Genomic DNA Purification Mini Spin Kit, as per the manufacturer’s instructions. DNA fragment detection was performed as previously published [3]. Briefly, a mixture of DNA, TdT, and dATP was prepared and incubated at 37□ for 30 minutes, followed by an additional incubation at 75□ for 20 minutes. Finally, Endo IV and the probe were added to the system. The fluorescence intensity was measured immediately using the real-time PCR instrument (RRID: SCR_018018).

#### 1.8 Cellular Thermal Shift Assay (CETSA)

After freeze-thawing the GBM cells twice using liquid nitrogen to get the whole protein, divide the protein solution into two equal parts. Incubate each part with DMSO or Z5 at room temperature for 1 hour. After incubation, aliquot the protein solution and incubate at different temperatures for three minutes. Following this, centrifuge the samples at 12000g, collect the supernatants, and perform immunoblotting analysis.

#### 1.9 Drug Affinity Responsive Target Stability Assay (DARTS)

DARTS was performed as previously published^2^. Briefly, after lysing GBM cells with RIPA buffer, divide the protein solution into five equal parts, with each portion being 90 μL. Measure the protein concentration using the BCA kit. Meanwhile, incubate each portion of the protein solution with DMSO or different concentrations of Z5 at room temperature for 15 minutes. Based on the protein concentration, add protease solution at a ratio of 1:700 (enzyme: protein) and incubate at room temperature for 10 minutes. Subsequently, stop the reaction by adding a protease inhibitor cocktail and SDS-PAGE loading buffer followed by immunoblotting.

#### 1.10 Surface plasmon resonance (SPR)

A Biacore 1K instrument was used to measure the binding kinetics of the human EGFR kinase domain (EGFR residues 704-1016) and human WEE1 kinase domain (WEE1 residues 293-568) to Z5. Measurements were performed at 25□. Proteins were dissolved in PBS, pH7.4. The proteins were immobilized on the CM5 sensor chip (Cytiva, 29149603, Shanghai, China) using the amine-coupling method according to standard protocols. Data were processed using standard double-referencing and fit to a 1:1 binding model using Biacore 1K Evaluation software. The association rate (K_on_, M^−1^□s^−1^), dissociation rate (K_off_, s^−1^), estimate of error (SE), Chi-Square (Chi^2^), and statistic and maximum response (R_max_) and response units (RU) were determined. The equilibrium dissociation constant (K_d_) was calculated from the relationship *K_D_*□=□K_off_/K_on_ (M). GraphPad Prism 9.0 was used for image processing.

#### 1.11 Transfection of siRNA and plasmids

U87-MG and U251-MG cell lines were transfected with siRNA using siRNA-Mate (GenePharma, Suzhou, China). Transfections were carried out following the siRNA-Mate protocol. The siRNA sequences were shown in **Supplementary Table S3**. The pcdna3.1 (Vehicle), EGFR WT and EGFR E762V plasmids were purchased from Gene Create (Wuhan, China). The plasmids were transfected into the HEK293T cells using Lipofectamine 3000 (Thermo Scientific, USA) according to the manufacturer’s instructions. The plasmid sequences were shown in **Supplementary Table S4**.

#### 1.12 Limited Proteolysis–Mass Spectrometry (LiP-MS)

Recombinant EGFR-kinase domain (EGFR residues 704-1016) were incubated with 100 μM of Z5 at 25°C for 15 minutes; Control Group: Recombinant EGFR-kinase domain were incubated with an equal volume of DMSO at 25°C for 15 minutes. After incubation, Proteinase K was added to the protein solutions (enzyme-to-protein ratio by weight is 1:100), and the mixture is digested at 25°C for 3 minutes, followed by immediate heat treatment at 98°C for 5 minutes to stop the enzymatic reaction. The samples, post-Proteinase K digestion, were cooled to room temperature, then an equal volume of 2% SDC (prepared in 20 mM Tris-HCl buffer) is added, adjusting pH to 7-8.5 with ammonium bicarbonate. The samples were heated at 98°C for 5 minutes, cooled to room temperature, and then 5 μL of 0.1 M TCEP and 5 μL of 0.4 M chloroacetamide were added, reacting in the dark at 45°C with shaking at 1500 rpm for 5 minutes. After cooling the samples to room temperature, trypsin (Promega, Madison, WI) was added at an enzyme-to-sample ratio (by weight) of 1:50 and incubated overnight at 37°C for proteolysis. An appropriate amount of formic acid was added to achieve a final concentration of 1.5%, mixed thoroughly, and centrifuged at 16000g for 5 minutes. The supernatant was collected, desalted using a C18 desalting column, and dried under vacuum. Each sample had been resuspended in 30 μL of Solvent A (A: 0.1% formic acid in water) to create a suspension. Then, 9 μL of the suspension was mixed with 1 μL of 10× iRT peptide mix. After mixing thoroughly, the samples were subjected to separation by nano-Liquid Chromatography (nano-LC). The separated peptides were analyzed by online electrospray tandem mass spectrometry (MS/MS). The differential peptides identified by LIP-MS can be found in **Supplementary Table S5**.

#### 1.13 qPCR

Total RNA was extracted from cells with Trizol and was transcribed to cDNA using HiScript QRT SuperMix reverse transcriptase (Vazyme, R223–01, Nanjing, China). qPCR was then performed on ABI QuantStudio 5 (Thermo Fisher Scientific, USA, RRID: SCR_018018) using SYBR Green qPCR Mix (Vazyme Biotech Co.,Ltd, China). Relative mRNA levels were normalized to GAPDH. Primer sequences were listed in **Supplementary Table S6**.

#### 1.14 Cell cycle detection assay

Following drug treatment, cells were harvested by centrifugation at 300 × g for 5 min, and the supernatant was carefully removed. The cell pellet was resuspended in phosphate-buffered saline (PBS) and re-centrifuged under the same conditions to wash. Subsequently, cells were fixed by slow addition of ice-cold 70% ethanol while vortexing, and stored at 4□°C overnight.

On the following day, fixed cells were pelleted by centrifugation and washed once with PBS. For combined phospho-histone H3 (PHH3) and propidium iodide (PI) staining, cells were first incubated with primary anti-PHH3 antibody at room temperature for 1 hour with gentle agitation. Following three washes with PBS, cells were incubated with Alexa Fluor® 488-conjugated anti-mouse IgG secondary antibody (dilution 1:200) in the dark at 4□°C for 1 hour. After three additional PBS washes, cells were resuspended in staining buffer containing 0.05% Triton X-100, 50□μg/mL PI, and 100□μg/mL RNase A, and incubated at 37□°C for 30 minutes in the dark. Finally, cells were washed three times with PBS before analysis using a flow cytometer.

For PI single staining, an identical procedure was followed, omitting the PHH3 antibody and secondary antibody incubations. Instead, cells were directly subjected to permeabilization and staining with PI/RNase solution under the same conditions as described above.

#### 1.15 Immunofluorescence assay

Immunofluorescence analysis was performed as previously published^4^. All images were captured using a confocal laser scanning microscope (Nikon AX/AX R with NSPARC, Japan). The primary antibodies used are listed in **Supplementary Table S2.** For GBM cell lines, secondary antibodies were anti-rabbit IgH (H+L) (Alexa Fluor 647 Conjugate) (RRID : AB_10693544) and anti-mouse IgH (H+L) (Alexa Fluor 488 Conjugate) (RRID: AB_10694704). For GBM patient tissue, secondary antibodies were anti-rabbit IgH (H+L) (Alexa Fluor 488 Conjugate) (RRID : AB_1904025) and anti-mouse IgH (H+L) (Alexa Fluor 488 Conjugate) (RRID: AB_1904023).

#### 1.16 Co-Immunoprecipitation assay

U87-MG and U251-MG cells were treated with Z5 for 15 min, followed by lysis in RIPA buffer containing protease and phosphatase inhibitors. Lysates were incubated on ice for 15 min and cleared by centrifugation (12,000 × g, 10 min, 4□°C). An aliquot of each lysate was reserved as whole-cell lysate (WCL) input. The remaining lysate was incubated with anti-EGFR antibody (Santa Cruz Biotechnology) at 4□°C for 1 h with rotation. Pre-washed Protein A/G PLUS-Agarose beads (Santa Cruz Biotechnology) were then added, and the mixture was incubated overnight at 4□°C. The beads were washed three times with ice-cold RIPA buffer, resuspended in 1× SDS loading buffer, and boiled at 95□°C for 10 min to elute bound proteins. Samples were analyzed by SDS-PAGE and immunoblotting with specific antibodies.

#### 1.17 Clinical Bioinformatics Analysis of WEE1

Bioinformatic analyses of WEE1 expression and its correlation with patient survival in GBM were conducted using data from the Gliovis database.

## 2. Supplementary Figures and Legends

**Supplementary Figure 1.**
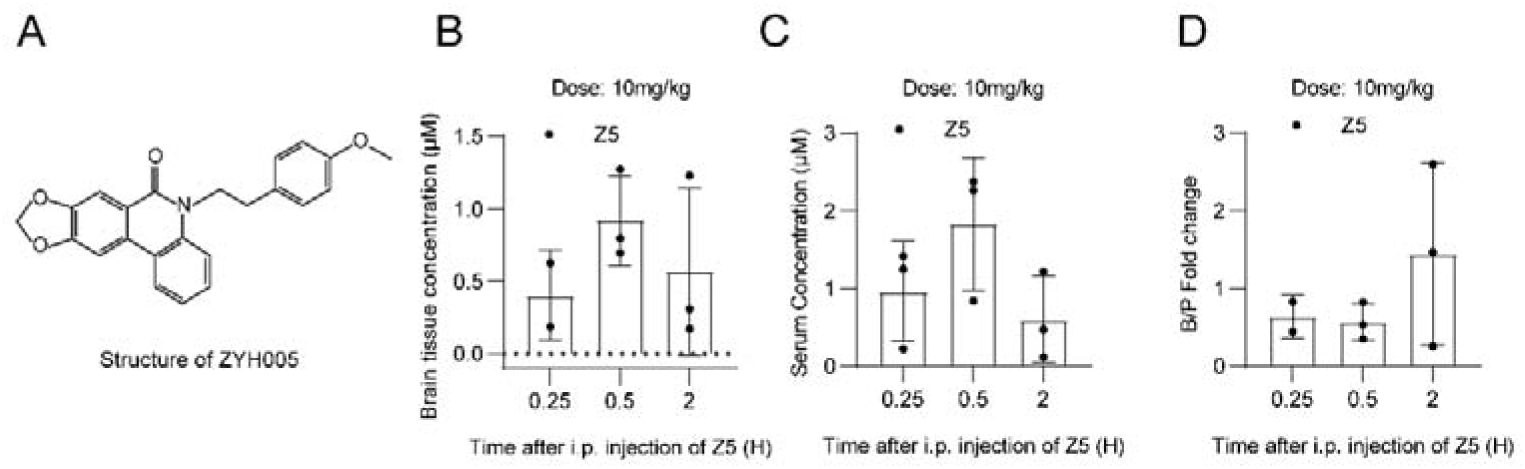
Structure and permeability of Z5. (A) Chemical structure of Z5. (B) Detection of intracranial drug concentrations at different time points after intraperitoneal injection of Z5 using LC-MS/MS (n=3). (C) Detection of plasma drug concentrations at different time points after intraperitoneal injection of Z5 using LC-MS/MS (n=3). (D) Ratio of intracranial drug concentration to plasma drug concentration at different time points after intraperitoneal injection of Z5 (n=3).

**Supplementary Figure 2.**
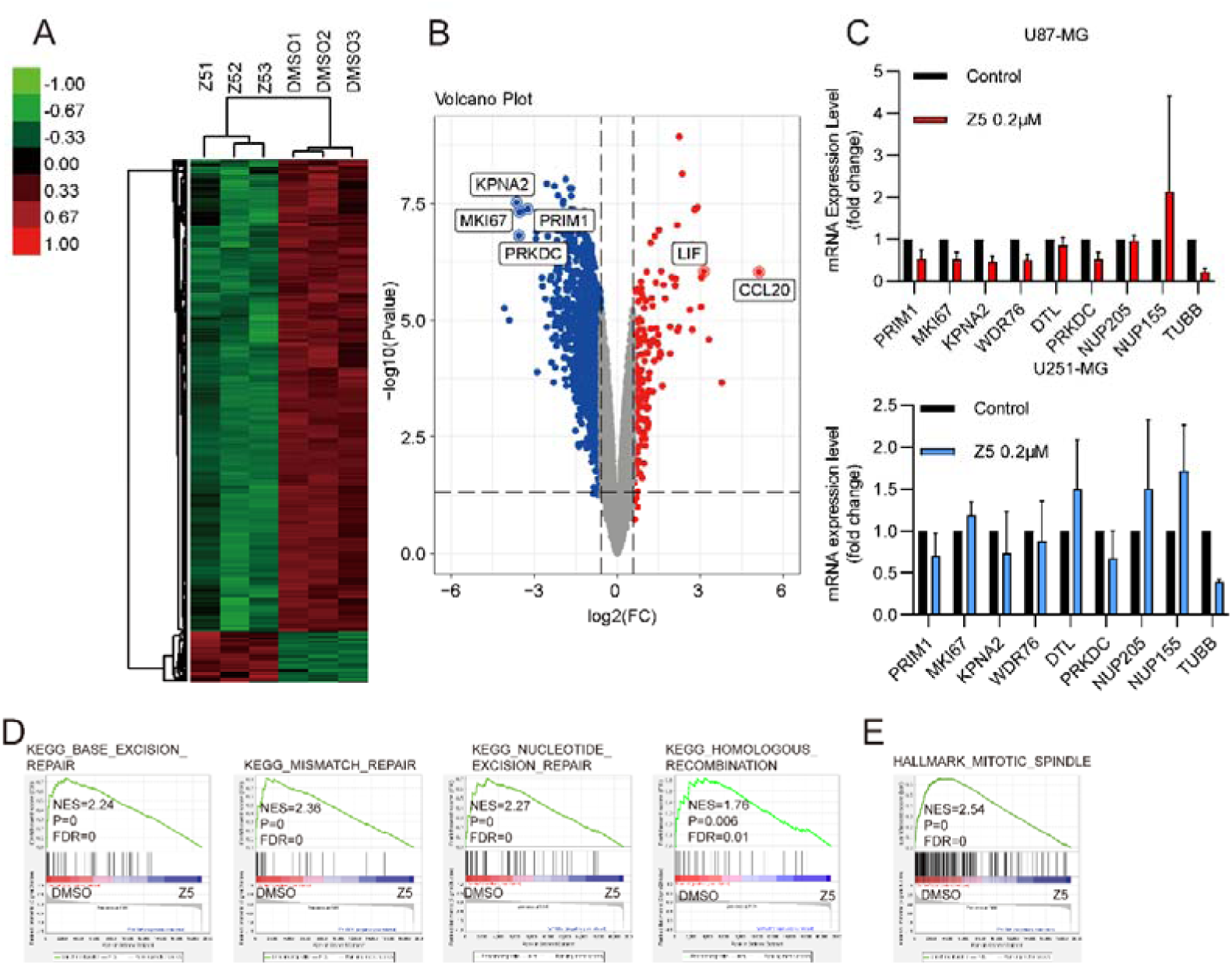
Microarray data and GSEA analysis of Z5 treated GBM cells. (A) Heatmap of microarray data from U87-MG cells treated with DMSO or Z5. (B) Volcano plot showing differentially expressed genes in U87-MG cells after Z5 treatment (|log□ fold change| > 1, adjusted *p* < 0.05). (C) qPCR validation of selected differentially expressed genes in GBM cell lines. (D) GSEA analysis of base excision repair, mismatch repair, nucleotide excision repair and homologous recombination pathways. (E) GSEA analysis of mitotic spindle pathways.

**Supplementary Figure 3.**
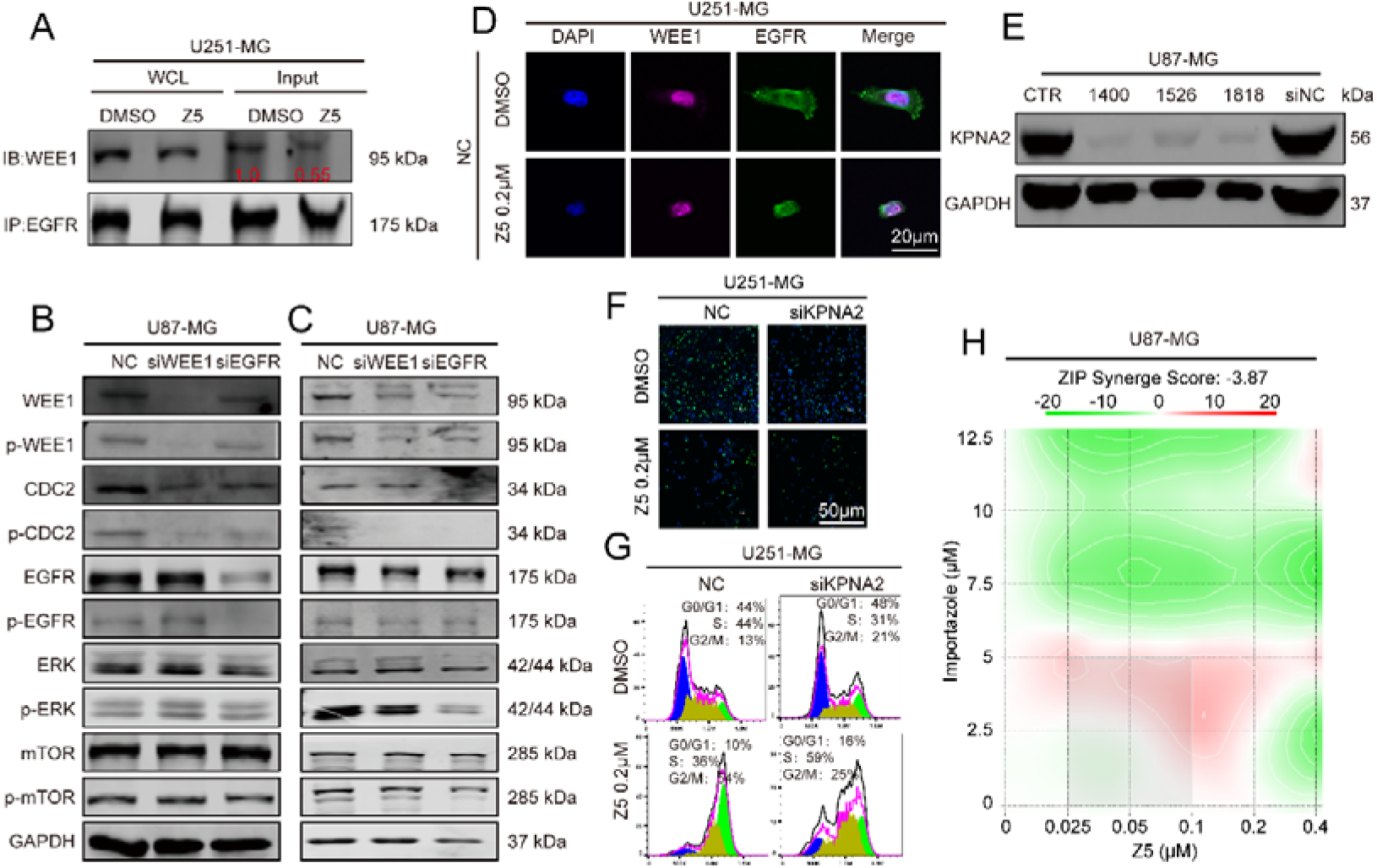
Z5 disrupt the nuclear EGFR-WEE1 axis. (A) Co-IP analysis of EGFR–WEE1 interaction in U251-MG cells after 15 min DMSO or Z5 treatment in U251-MG cells. (B-C) Immunoblot analysis of WEE1, p-WEE1, CDC2, p-CDC2, EGFR, p-EGFR, ERK, p-ERK, mTOR, and p-mTOR in U87-MG cells following WEE1 or EGFR knockdown, or treatment with MK1775 (4 μM) and AZD9291 (8 μM). (D)Immunofluorescence (IF) staining was used to analyze the localization of EGFR and WEE1 in U251-MG cells. (E) Efficiency of KPNA2 silencing by siRNA transfection was assessed via immunoblot. (F) Anti-proliferative effect of Z5 in NC and si-KPNA2 GBM cells was evaluated by EdU staining. (G) Cell cycle distribution in NC and si-KPNA2 U251-MG cells was analyzed by flow cytometry after 24 h Z5 treatment. (H)ZIP synergy score analysis of the combinatorial effect between Z5 and the KPNB1 inhibitor importazole.

**Supplementary Figure 4.**
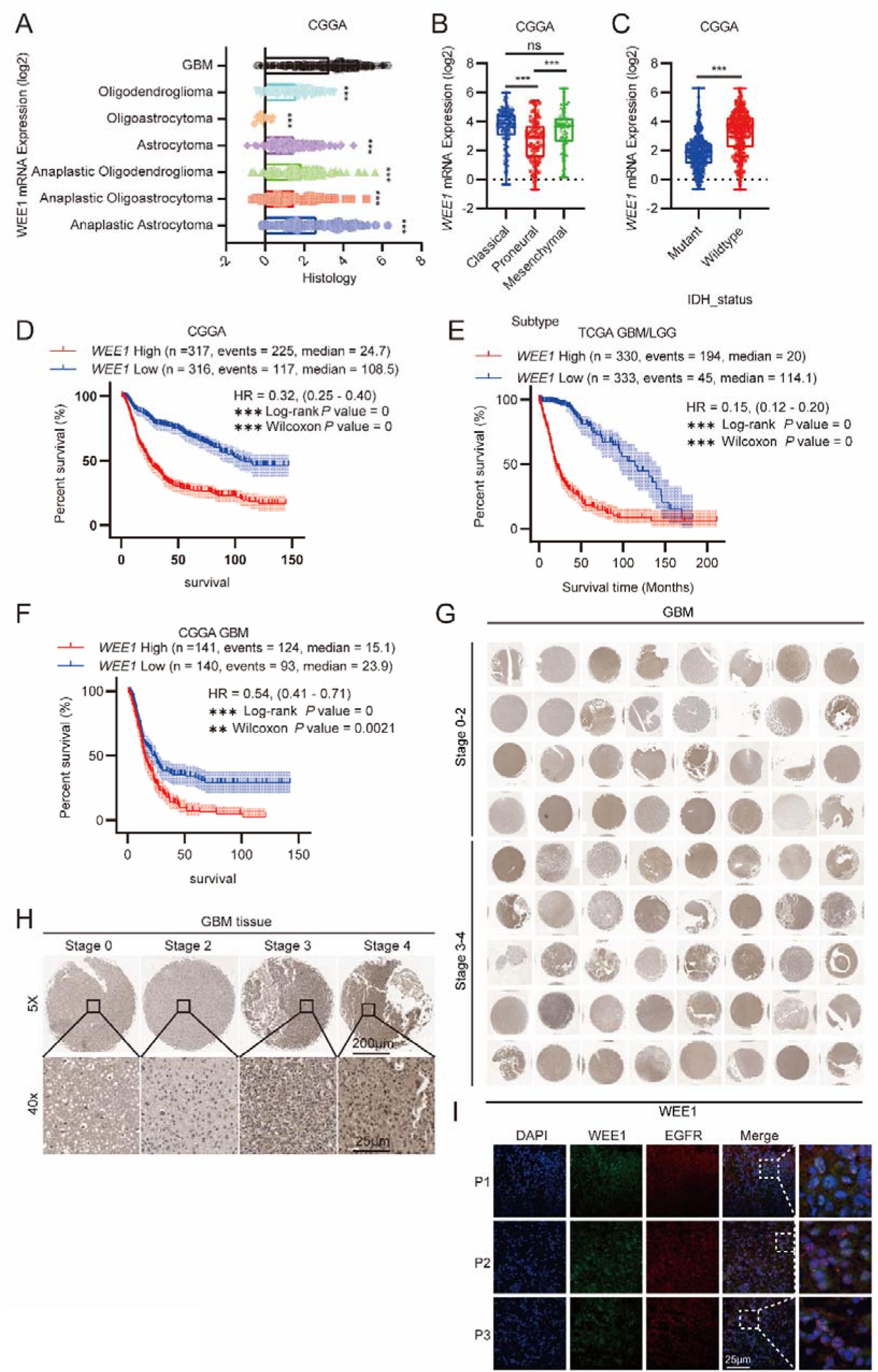
WEE1 expression correlates with poor survival and EGFR levels. (A) WEE1 expression across glioma subtypes in the CGGA cohort. (B) WEE1 expression across GBM subtypes in the CGGA database. (C) WEE1 expression in IDH-mutant and IDH-wildtype GBM. Survival analysis based on WEE1 expression in (D) glioma patients (CGGA), (E) GBM/LGG patients (TCGA), and (F) GBM patients (CGGA). (G) Tissue microarray (TMA) of GBM samples. (H) Representative images of WEE1 immunohistochemistry on the TMA are shown (original magnification: upper: 5x; lower: 40x). (I) IF staining images of tumor tissues from GBM patients.

**Supplementary Figure 5.**
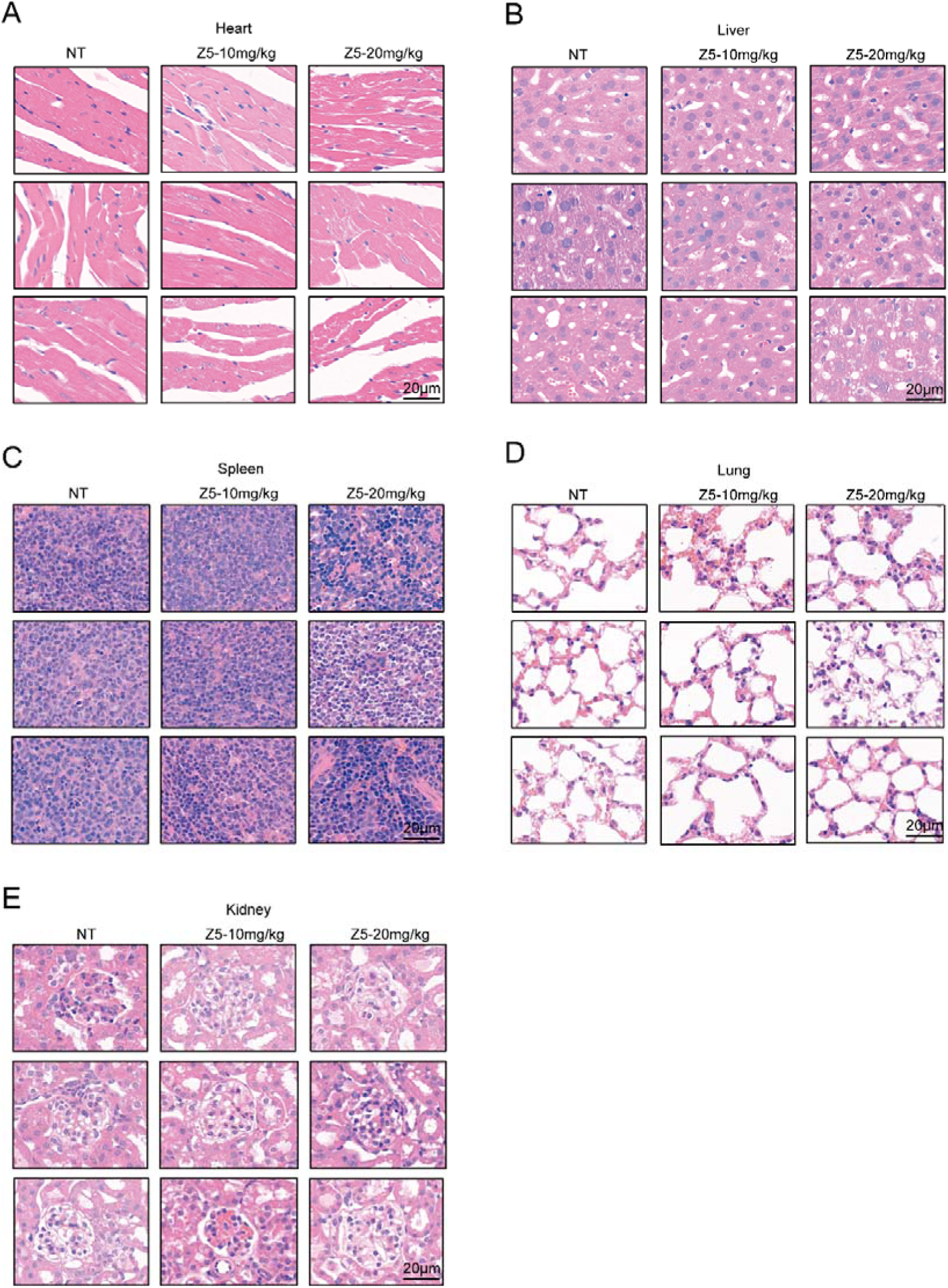
Histological Evaluation of Organ Toxicity by Z5 Using H&E Staining. (A) H&E-stained sections of the heart. (B) H&E-stained sections of the liver. (C) H&E-stained sections of the spleen. (D) H&E-stained sections of the lung. (E) H&E-stained sections of the kidney.

**Supplementary Figure S6.**
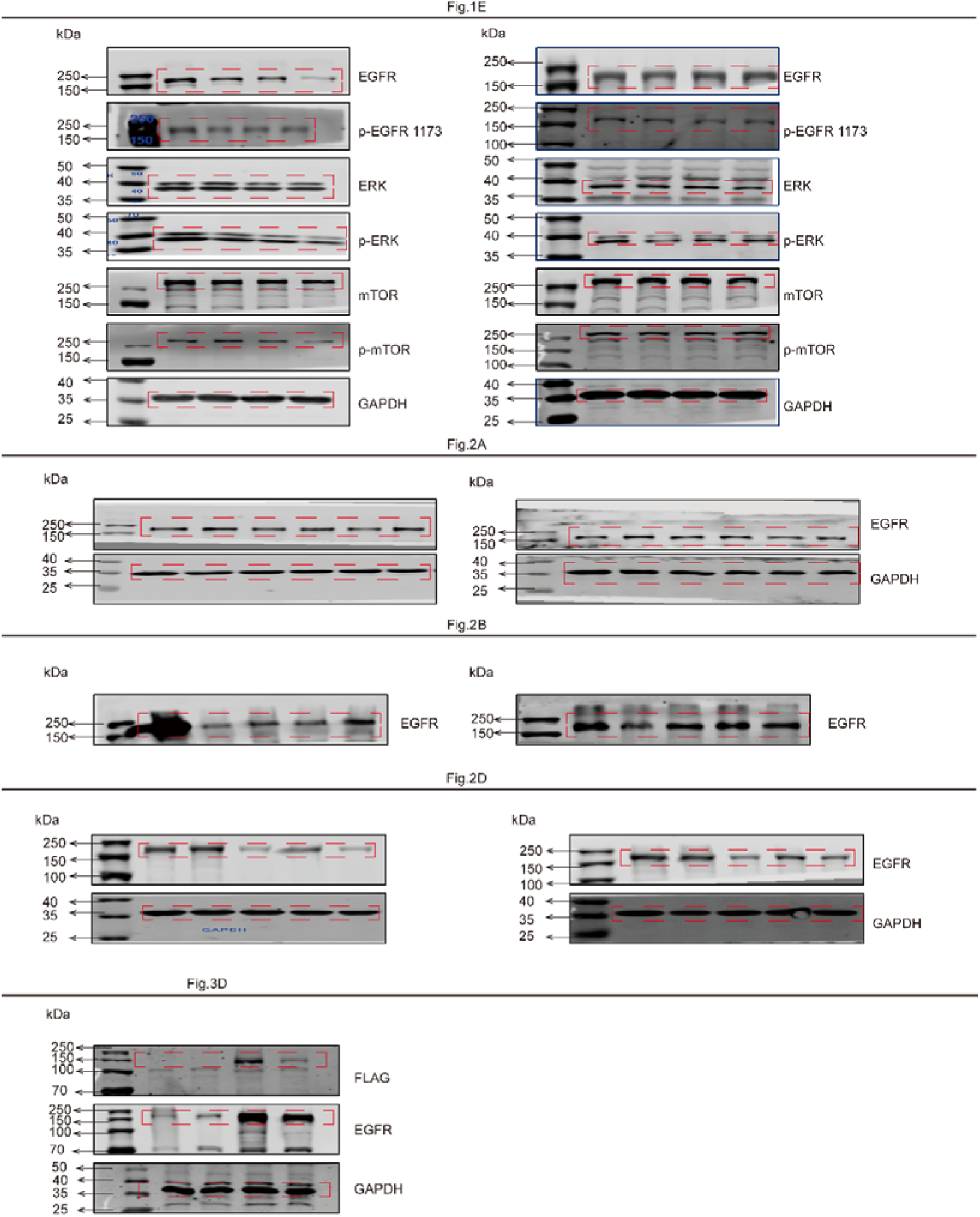
Complete membrane images of immunoblotting results shown in Fig. 1E, 2A, 2B, 2D and 3D.

**Supplementary Figure S7.**
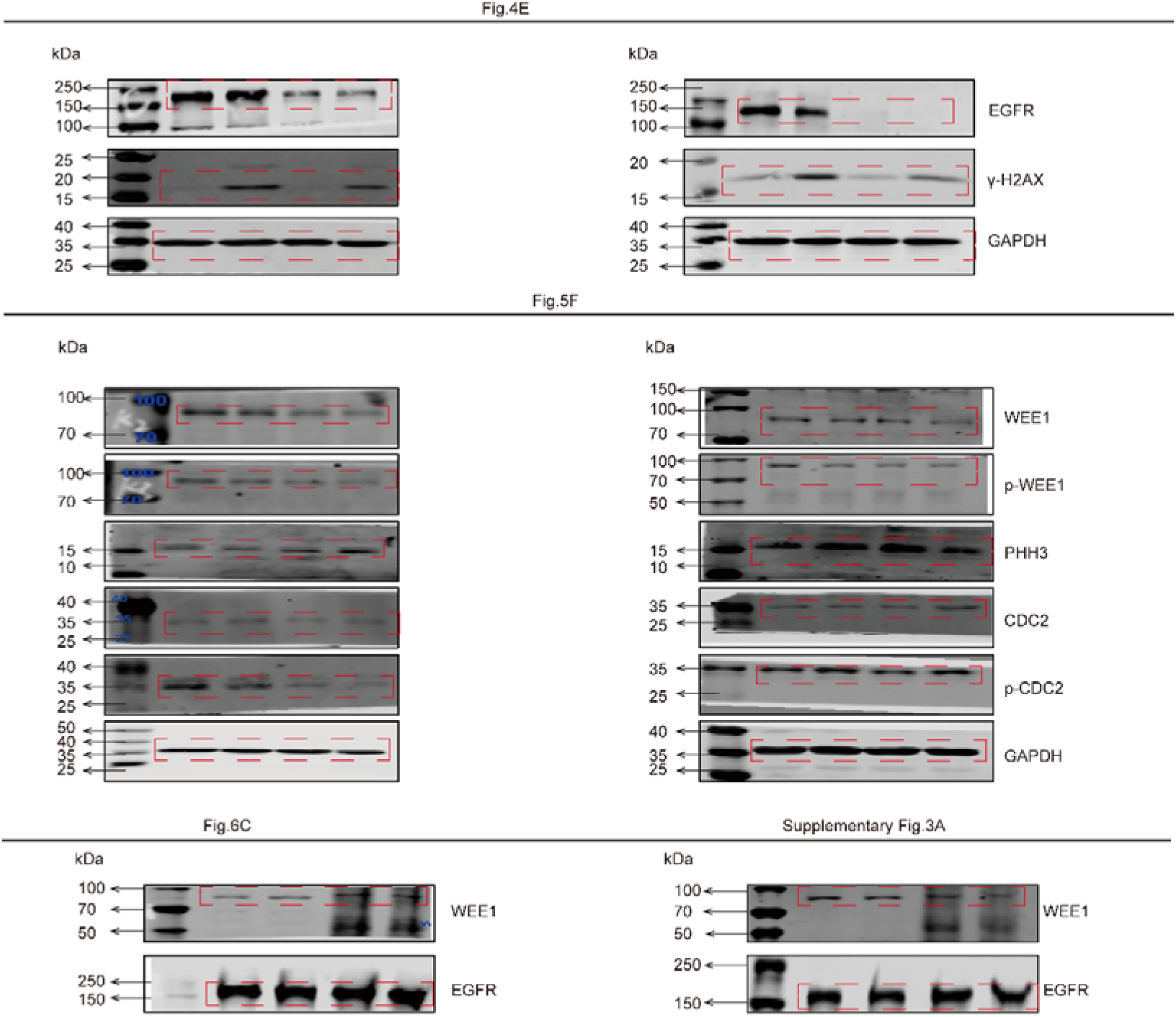
Complete membrane images of immunoblotting results shown in Fig. 4E, 5F, 6C and Supplementary Fig.3A.

**Supplementary Figure S8.**
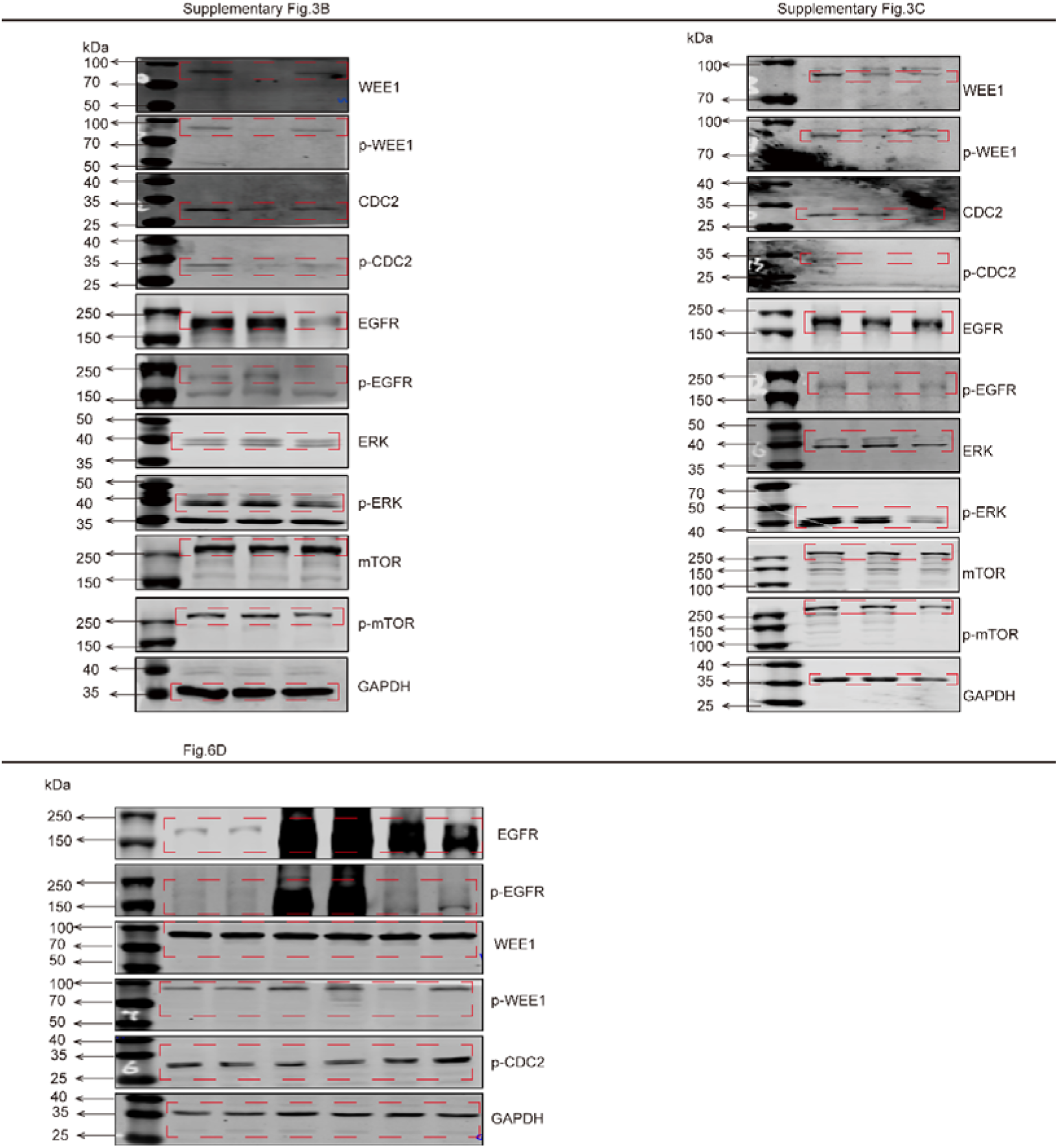
Complete membrane images of immunoblotting results shown in Fig.6D and Supplementary Fig. 3B, 3C.

**Supplementary Figure S9.**
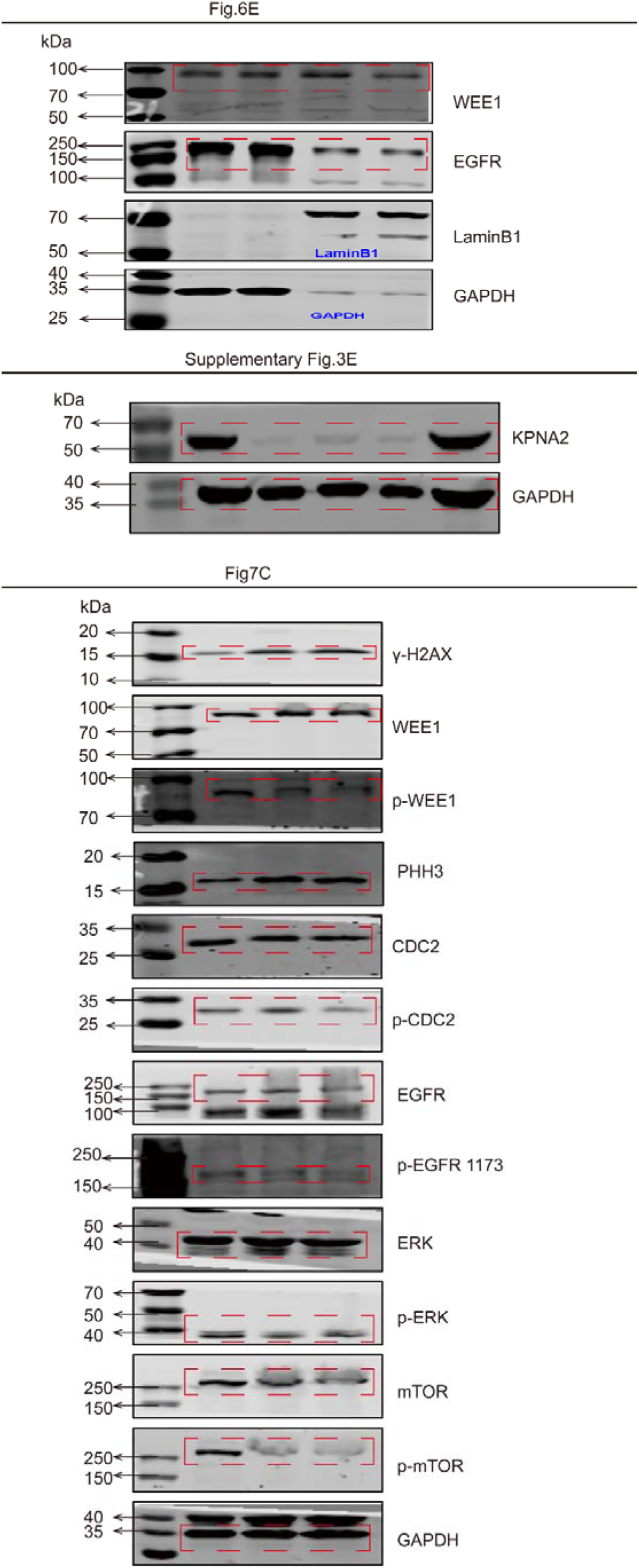
Complete membrane images of immunoblotting results shown in Fig. 6E, 7C and Supplementary Fig. 3E.

